# Modeling COVID-19 Nonpharmaceutical Interventions: Exploring periodic NPI strategies

**DOI:** 10.1101/2021.02.28.21252642

**Authors:** Raffaele Vardavas, Pedro Nascimento de Lima, Lawrence Baker

## Abstract

We developed a COVID-19 transmission model used as part of RAND’s web-based COVID-19 decision support tool that compares the effects of nonpharmaceutical public health interventions (NPIs) on health and economic outcomes. An interdisciplinary approach informed the selection and use of multiple NPIs, combining quantitative modeling of the health/economic impacts of interventions with qualitative assessments of other important considerations (e.g., cost, ease of implementation, equity). This paper provides further details of our model, describes extensions, presents sensitivity analyses, and analyzes strategies that periodically switch between a base NPI level and a higher NPI level. We find that a periodic strategy, if implemented with perfect compliance, could have produced similar health outcomes as static strategies but might have produced better outcomes when considering other measures of social welfare. Our findings suggest that there are opportunities to shape the tradeoffs between economic and health outcomes by carefully evaluating a more comprehensive range of reopening policies.

## 1 Introduction

Coronavirus disease 2019 (COVID-19) is unprecedented in terms of scale and speed, affecting millions worldwide. Until recently, vaccines and effective treatments for COVID-19 were unavailable. National leaders have had to take extraordinary measures to mitigate the virus’s spread and prevent health care systems from being overwhelmed. Policymakers have implemented a range of nonpharmaceutical public health interventions (NPIs). These interventions include partial closings (e.g., schools and non-essential businesses, prohibiting large gatherings, quarantining the most vulnerable) and complete lockdown (e.g., placing all residents under stay-at-home orders). The goal of NPIs is to delay and reduce the peak number of cases per day, reduce pressure on health services, and allow time for vaccines to be distributions [1]. If NPIs are relaxed too soon a new wave of infections may occcur. However, NPIs have wide-ranging effects on the health, economy, and social well-being of populations, which has led to growing pandemic fatigue and a decline in adherence to NPIs since they were first initiated [2, 3]. Decision-makers are faced with tough decisions, such as how to sequence, relax, and possibly reinstate mitigation measures. Exacerbating these decisions are significant uncertainties, including new variants and behavioral responses to extended interventions.

Mathematical and simulation models of COVID-19 transmission dynamics are invaluable tools to help decision-makers forecast and compare intervention outcomes, predict the timing of peaks in cases and deaths, medical supply needs, and if and when we should expect additional waves. They enable projection and comparison of population-level outcomes over hypothetical scenarios. Model outcomes include the incidence and prevalence of the infection over time and for different population groups. The hypothetical scenarios can consist of the impact of varying pharmaceutical and nonpharmaceutical public health interventions, distributing vaccines, and the emergence of new strains.

We developed a COVID-19 transmission Population-Based Model (PBM) used as part of a web-based COVID-19 decision support tool that compares the effects of different nonpharmaceutical public health interventions (NPIs) on health and economic outcomes. An interdisciplinary approach informed the selection NPI portfolios, combining quantitative modeling of the health/economic impacts with qualitative assessments of cost, ease of implementation and equity. An in-depth description of our approach was previously published as a RAND report describing how the PBM, the economic model, and a systematic assessment of NPIs informed the web-tool [4].

We expanded our original model [4] to account additional uncertainties and consider an expanded set of NPI strategies. In this paper, we consider periodic NPI strategies. Recent research has demonstrated that high-frequency periodic NPIs [5] have the potential to mitigate COVID-19 resurgences while providing more predictability and alleviating the damaging effects on economic activity and social well-being. We use our updated model to explore if a periodic strategy could have provided benefits compared to fixed strategies that would keep *R*_*t*_ close to one. We find that a periodic strategy can dominated fixed strategies, improving both health and days spent under restrictions.

The paper is structured as follows. First, we provide an overview of our model structure. Then, we briefly analyze a set of illustrative scenarios, including periodic and fixed strategies, identifying if the periodic strategies used by other modelers modelers [5] produce similar results in our model. Finally, we provide detailed information on our mathematical model and present sensitivity analyses.

## 2 Model Overview

Theory-based epidemiological models use a theoretical understanding of biological and social processes to represent a disease’s clinical and epidemiological course. The most typical model considers the population in four different disease states: susceptible, exposed, infected, and Removed (SEIR). Our PBM incorporates several extensions to the SEIR model of disease transmission. It is formulated deterministically by coupled ODEs and integrated numerically by solvers for stiff problems [6–9]. We extend the SEIR framework to better describe COVID-19 transmission by adding additional disease states and considering population strata based on age and chronic conditions. The PBM models the effects of different NPIs on health outcomes, from partial closings to complete lockdown. Unlike many other COVID-19 models, we simulate the impact of NPIs on different mixing modes (such as home, school, and work) separately, allowing us to model various interventions flexibly. Our PBM also includes population strata and specify mixing pattern heterogeneities across the population strata and for each mode. These heterogeneities included in our model allow us to set the NPI more specifically, with mixing rates reduced deferentially by mixing mode. Our model is designed to help policymakers understand the effects of NPIs, weigh tradeoffs among them, make decisions about which NPIs to implement, and estimate how long-term interventions should be enforced to control the virus.

Figure 1 shows a simplified illustration of the disease states included in the first basic version of our PBM. The model includes a pre-symptomatic highly infectious state, which leads to either an asymptomatic state or a state with mild symptoms, a fraction of which continue to severe disease. Most of those who develop severe symptoms are hospitalized. Non-hospitalized severely-symptomatic either recover or die.

**Figure 1:**
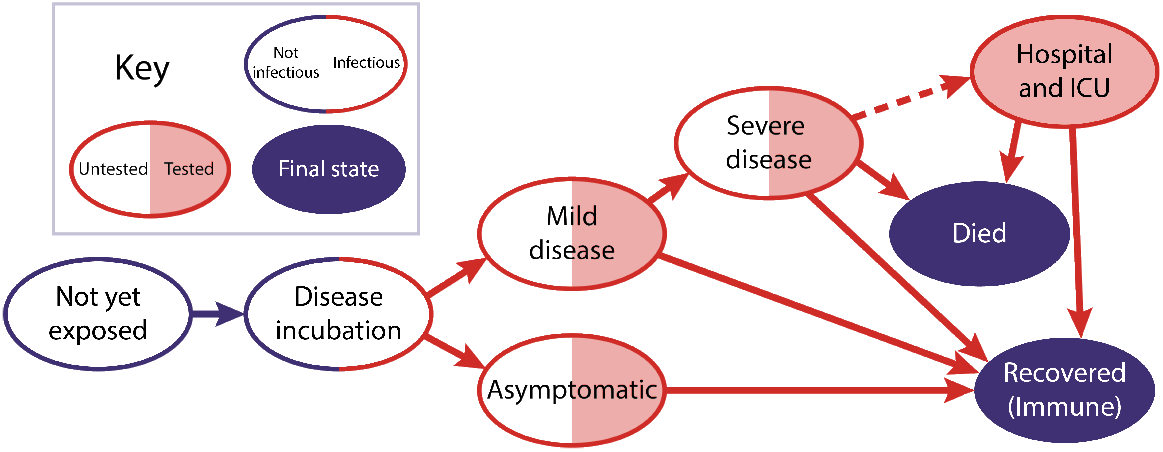
COVID-19 PBM disease states

The hospitalized state includes compartments for both the main hospital and the ICU, where individuals are admitted if they develop critical symptoms. Capacities can be set for the hospital and ICU beyond which no additional patients can be added. At each of the infectious states, individuals can be tested for COVID-19. Each compartment is composed of ten population strata, five age groups, and two health states (those with and without at least one chronic condition). These strata allow the model to simulate how the disease impacts different population groups, including differences in population size, mixing mortality rate, and the proportion who are asymptomatic. Disease progression rates are based on figures given in the literature and are sampled from distributions if uncertain. The force of infection (the rate that susceptible individuals become exposed) is characterized by how many infectious people are in each state, the transmissibility of each state, and levels of mixing. We estimate transmissibilities for each state based on biological and social factors. For instance, viral loads are highest early in the disease [10], so these states have higher biological transmissibility. We assume that those who receive positive tests or exhibit symptoms have lower social transmissibility because they take measures to limit the exposure of others. The total transmissibility of a state is the product of the biological and social transmissibilities, and the population-weighted sum of transmission is proportional to the number of new infections.

In our model, NPIs are portfolios of restrictions mandated at the state level, as described in table 1. The set of NPIs levels used by each state is characterized by a discrete set of intervention levels ranging from 1 (no intervention) to 6 (close schools, bars, restaurants, and nonessential businesses; and issue a shelter-in-place order for everyone but essential workers). Each intervention level is associated with mixing matrices that describe how strata interact with each other in six different settings: household, work, school, commercial, recreation, and other. Interventions are modeled as changing the level of mixing which occurs in each of these settings. For instance, closing schools reduces school and work mixing but increases home mixing. Given the specified model structure, the NPI time-series, and the mixing matrices, we calibrate our model for each state using deaths time series. Appendix A provides a detailed description of the mathematical formulation of our model.

**Table 1:**
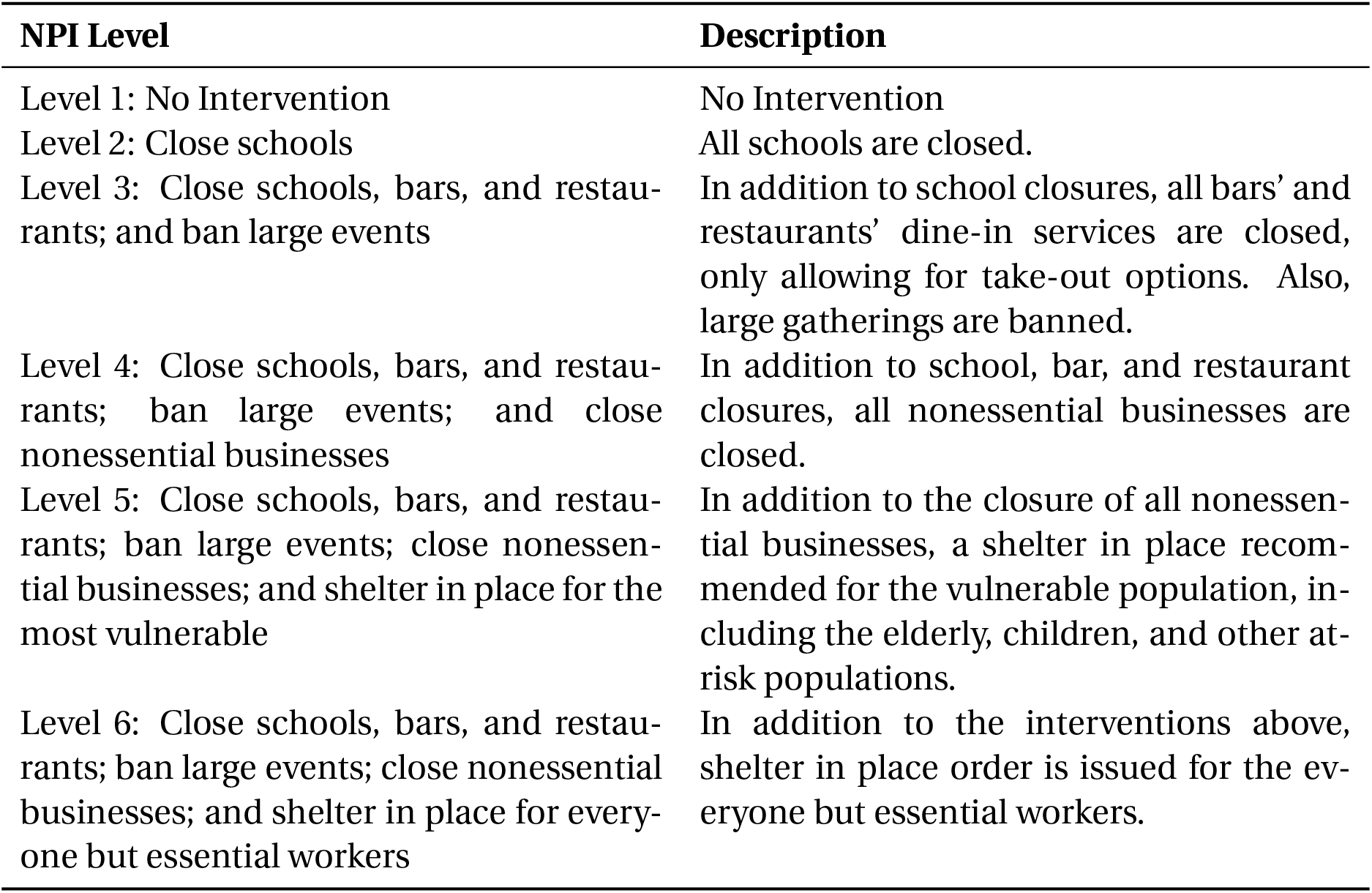
Nonpharmaceutical intervention levels.

## 3 Exploring Periodic NPI Strategies

In this section, we present an illustrative retrospective analysis of policies that can be tested with the model, using the state of California as an example. These analyses and findings do not constitute predictions. Yet, they illustrate that alternative plausible NPI strategies could have produced improved outcomes during 2020, in the absence of vaccines. The purpose of this analysis is two-fold. First, it demonstrates how our model can be used to trace many-objective trade-off curves to support the analysis of reopening strategies. Second, this analysis demonstrates that a periodic switching of NPIs might have placed society in a better position in these trade-off curves - that is, it could have represented a pareto-improvement.

This section explores two types of strategies that could have been followed to manage NPIs in the year 2020. The first set of strategies are “fixed” NPI levels. This type of strategy holds the NPI level constant over time. Although this strategy has not been followed in California explicitly, the NPI mandates imposed in California are best approximated in our model by setting the NPI level to three. This NPI level was stable between July of 2020 through the end of the year, and this represents our baseline scenario and is the scenario used to calibrate our model.

Fixed NPIs are not, however, the only way to control the pandemic. Alternatively, one could use periodic strategies to curb transmission [5]. A periodic NPI can represent a strategy wherein society goes into more severe periods of NPIs then relaxes to lower levels of stringency. This strategy’s rationale is that those newly infected during the relaxed periods would take a few days before becoming infectious themselves. The enforcement of stringent NPIs would then limit the time and possibilities for the virus to spread further from these infectious individuals during the time before they either recover or are hospitalized. This periodical switching would systematically reduce transmissions and force the dynamics of the epidemics to be controlled by the strategy’s frequency.

In essence, a periodic strategy uses the natural timescales of disease progression and infectivity to induce a synchronization phase that helps align the times when people are more likely to be infected together, allowing for the social distancing NPIs to be more effective. An example of a similar strategy includes schools adopting a hybrid learning model wherein students go to school every other week. Similarly, restaurants could open for indoor dining periodically. In absence of vaccines, such policies may be desirable as they would provide stability, regularity, and increased predictability for businesses to plan against. The policies could, in principle, help suppress the transmission of the virus and simultaneously reduce uncertainties in economic activity.

Figure 2 illustrates the dynamics of periodic and fixed NPI strategies. The fixed NPI strategies represented in the figure suggest that under the NPI level three, *R*_*t*_ closely followed one and increased towards the end of December, driven by our model’s seasonal effect. Because *R*_*t*_ was close to one in the model, a departure from the current NPI level would be expected to produce a significant departure from the *R*_*t*_ = 1. Therefore, a policy that reopens the state (F-1) would be expected to produce a spike in prevalence and subsequently in the number of deaths, and a more stringent, constant policy (F-5) would be expected to reduce the number of deaths.

**Figure 2:**
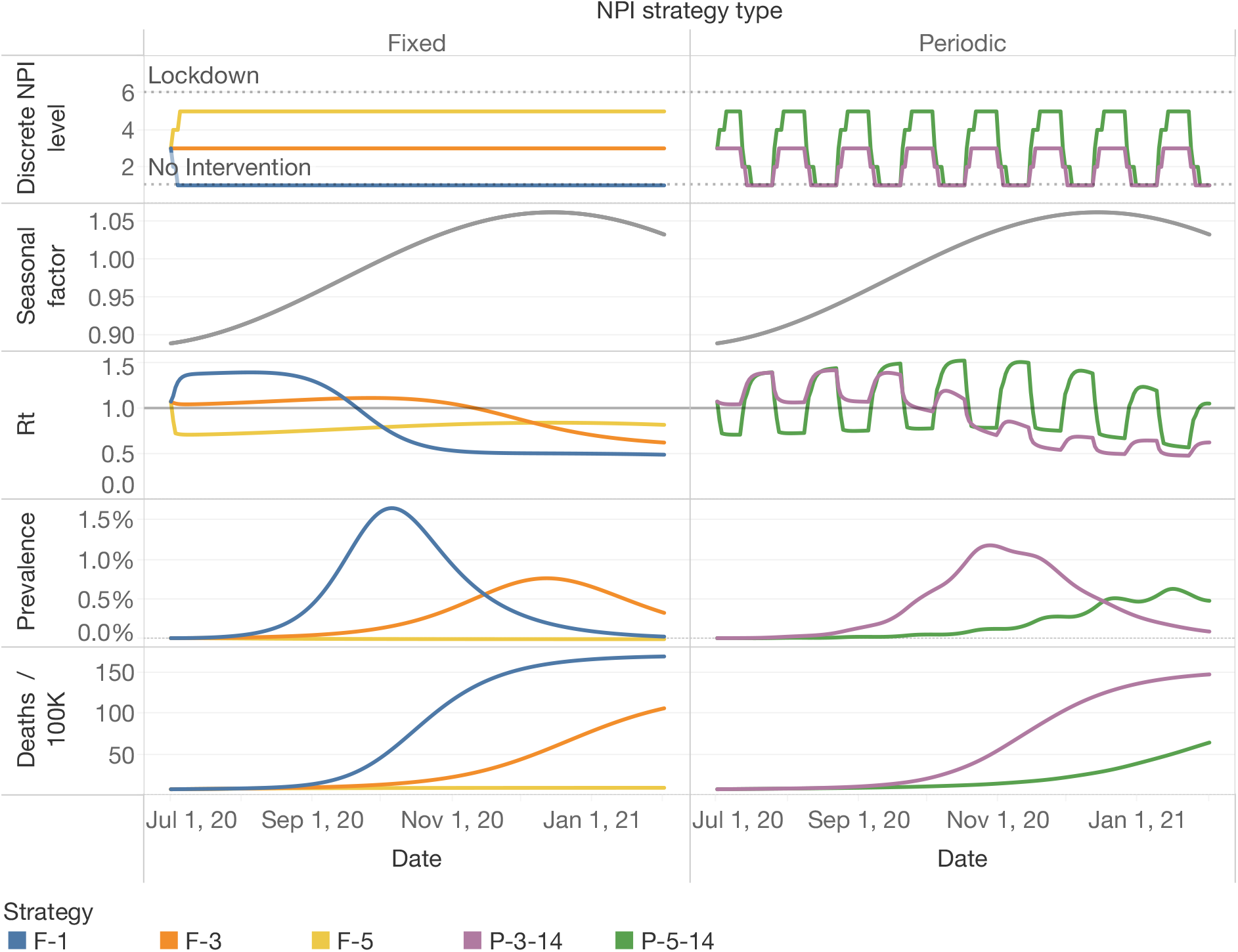
Model Dynamics with Fixed and Periodic NPIs. Fixed NPI strategies are coded with an “F” followed by the intervention level included in the NPI. Periodic NPI strategies are coded with a “P” followed by the maximum NPI level in the periodic strategy and the period in days. In that case, P-3-14 means that the state will switch between the NPI level 3 and 1 every 14 days.

As figure 2 shows, the periodic strategy P-5-14 switches between NPI levels 1 and 5 every two weeks. This switching causes *R*_*t*_ to oscillate such that prevalence does not increase unchecked. As a consequence, the number of deaths is controlled. The choice of two weeks is based on the typical timescale describing the disease progression for the majority of infected people. However, other choices for the periodicity could be explored.

When judging alternative strategies, policymakers often have to weigh multiple criteria to make decisions, so one needs to translate model outcomes to meaningful criteria. One criterion could be the number of days of school closures, which has been an important concern during the COVID-19 pandemic. However, the number of days of school closures does not distinguish scenarios in which non-essential businesses are closed for long periods, so other proxies for welfare loss are needed. One approach could be to use weights for each NPI level, such that those weights are proportional to the marginal daily welfare loss induced by each NPI level. If one defines those weights such that one day under lockdown is equivalent to one, and one day under no restrictions is set to zero, one can compute a proxy to social welfare that can be used to judge alternative strategies. Our weighted lockdown days metric corresponds to this criterion.

There are other plausible ways to compute NPI costs. NPIs arguably induce income loss. Our third metric uses the income loss under each NPI level estimated by a general equilibrium model [11] to account for the economic consequences of NPI restrictions. Although all these proxies are imperfect measures of social welfare loss induced by NPIs, our conclusions do not rely on their precision, but on the assumption that NPI costs are increasing in the level of restriction. This structural assumption allows us to illuminate trade-offs and reveal pareto-dominated strategies that rely on the structure of the epidemiological model^1^.

Figure 3 demonstrates that using a small set of alternative measures can support those decisions and reveal pareto-dominated strategies. Strategy F-3, our baseline strategy, is pareto-dominated by a wide range of strategies that oscillate between the NPI level of 5 and 1, using many periods, which is in line with prior research [5].

**Figure 3:**
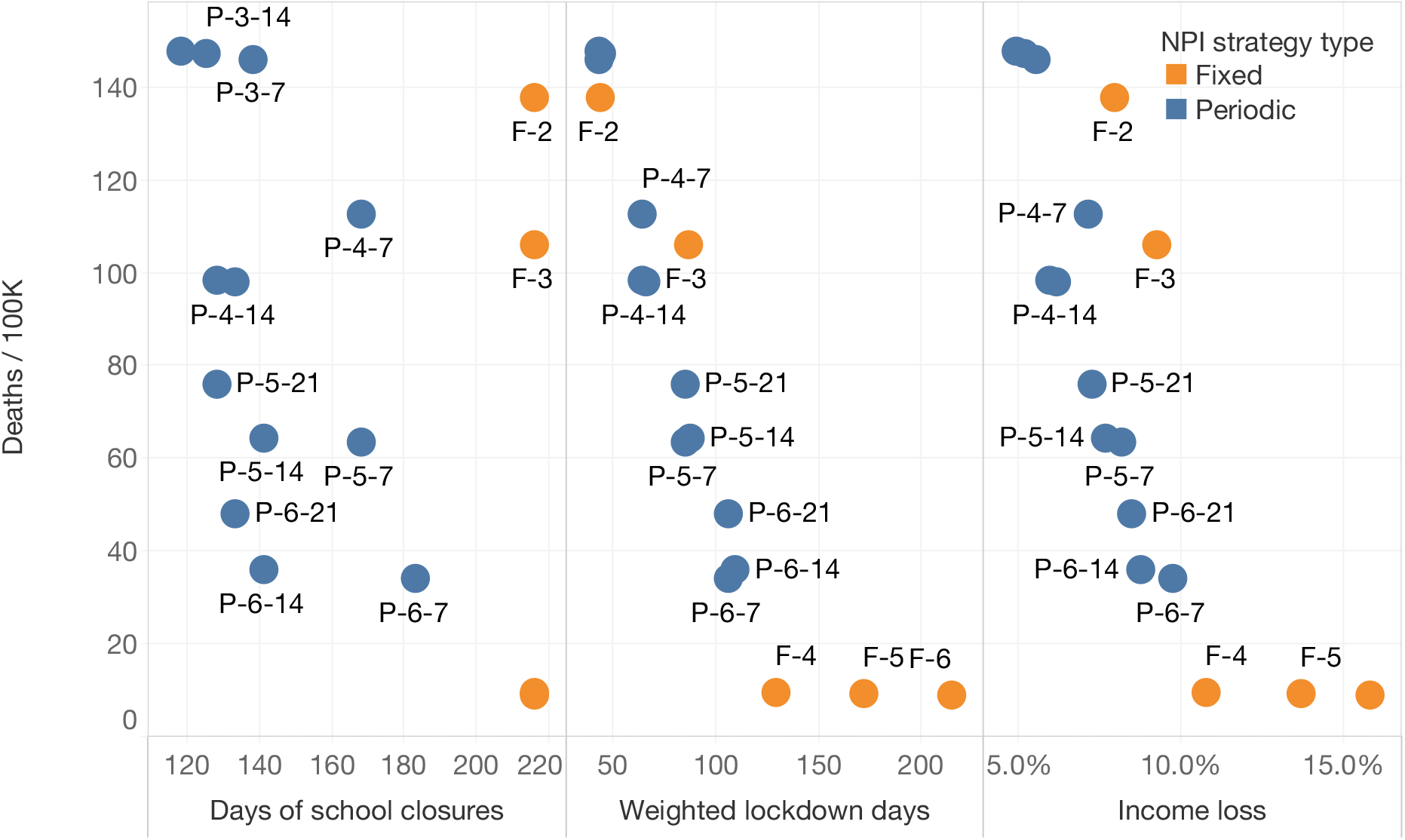
Tradeoff surface implied by alternative policies. The vertical axis presents the number of Deaths / 100k at the end of the simulation run (Feb. 2021) in California, and the horizontal axis contains several proxies that represent alternative criteria to evaluate the costs of NPIs. Across all these criteria, we find that periodic NPIs tend to dominate fixed NPIs. The plot demonstrates that strategies with fixed NPIs generally are dominated by periodic NPI strategies.

However, there are limitations to our analysis. First, we do not consider practicality: these periodic strategies might be regarded as unfeasible, impractical, or undesirable by policymakers and the public. This consideration is particularly important because the strength of the periodic NPIs relies on the ability to abruptly reduced mixing, which can only be achieved with a high level of compliance. Further, people are may to shift their mixing to the open periods reducing or even canceling the mitigation effects on transmission intended by the periodic NPI policy. Nevertheless, we use them in this paper as an example to illustrate that alternative NPIs strategies, if implemented with high levels of compliance, might pareto-dominate fixed strategies and might shift the trade-off surface among health and economic/social outcomes.

## 4 Conclusion

The scenarios presented in this paper serve the purpose to illustrate that alternative policies using periodic NPIs to manage the COVID-19 pandemic might have produced the same health outcomes while allowing essential activities, such as in-person education, to have happened in a controlled manner. While we do not advocate for any particular strategy, this brief analysis demonstrated that alternative NPI strategies have the potential to shift the trade-off curves among the relevant outcomes. Including social welfare loss measures induced by NPIs in analyses seeking to inform COVID-19 reopening decisions is essential. Only including health outcomes in those analyses leaves the task of weighing other concerns to the policymaker, who may or may not be able to do so consistently. Metrics of welfare loss induced by NPIs can be either derived directly from the model outcomes (e.g., days of school closures), or use a weighted sum based on the NPIs stringency level, potentially using economic models in our prior work[4]. Even if the analysis estimates are not precise or could become less precise with time, they can still be useful. As figure 3 demonstrated, if one ignores all the horizontal axes under the argument that those estimates might be imprecise, policymakers might not be properly informed that alternative policies dominate some policies. This statement and the pattern seen in the trade-off curves do not rely on the precision of economic estimates but on the theory-based epidemiological model structure.

The trade-off curves we present also should not be seen as static. Many other factors that have been held constant in our analysis might also shift this curve. Widespread adoption of high-quality masks, for example, would shift every point inwards, making society systematically better off. The emergence of new, more transmissible variant strains can shift the curve outwards. A more stringent strategy to eliminate community spread and prevent re-seeding (such as New Zealand’s strategy) can remove the health-economic trade-offs curve completely if successfully implemented. Adaptation measures to prevent transmission within schools would also shift this curve, strengthening periodic strategies even more attractive to allow in-person education. Moreover, the introduction of vaccines also shapes this trade-off curve over time. As vaccination roll-out advances, the marginal benefit of an additional day under stringent NPIs will decrease. Accounting for the uncertainties mentioned before and the vaccination dynamics will be essential to guide society to a new normal through a robust reopening plan.

## Data Availability

All modeling data is available upon request.

## Contributions

Raffaele Vardavas (Ph.D.) led the effort in conceptualizing and formulating the model structure and led the overall modeling effort. Pedro Nascimento de Lima (MSc) led the model implementation and model analysis effort. Lawrence Baker (MSc) led the effort in informing the model with data and parameter values found in the literature. All authors contributed equally to this research work and to this article.

## Acknowledgements

We wish to thank Drs. Jeanne Ringel, Jennifer Bouey, Courtney Gidengil and Carter Price at the RAND Corporation for their support in advising the authors in the model development and in finding parameter value estimates. We thank Drs. Robert Lempert, Carolyn Rutter at the RAND Corporation and Dr. Jonathan Ozik at the Argonne National Laboratory for their support in advising the authors in model calibration and policy exploration methods. We thank Dr. Chris Bauch, a professor in applied mathematics at the University of Waterloo for his constructive comments that significantly improved our model. This project was internally funded by RAND. We thank Drs. Michael Rich, Anita Chandra and Peter Hussey for supporting our work and securing funding. We also thank the National Cancer Institute (R21CA157571), and the National Institute of Allergies and Infectious Diseases (R01AI118705) for providing support in projects that led to preliminary work and ideas that motivated this project.

## Disclaimer

This report provides additional technical details of a peer-reviewed RAND report describing RAND’s COVID-19 Decision Support Tool for State and Local Policymakers. This report uses the simulation model to explore the dynamics of a retrospective counterfactual COVID-19 transmission mitigation policy for 2020 based on intermittent applications of NPIs. The results and conclusions drawn on how effective this policy design could have been if implemented based on our simulation model have not been peer-reviewed. Conclusions drawn from this working paper do not necessarily represent the opinions of the RAND Corporation.

## A The Mathematical Formulation of the Model

### A.1 Model disease states and progression

Individuals in our population are divided into fourteen key compartments listed in Table 2. We assume that individuals in the *P* and *I*_*A*_ compartments are fully asymptomatic and thus are unaware of being infectious. In our model, individuals in *I*_*Sm*_ have mild symptoms, including a dry cough and a fever, while those in *I*_*Ss*_ are assumed to have severe symptoms that include shortness of breath in addition to a dry cough and a fever. The sum of the population in all of the states gives the total population *N*. In our model we assume that each state variable gives the proportion of the population belonging to that state. Therefore, instead of tracking the dynamics of each compartment’s population sizes, we track the population densities. We express this as Σ_*X*_ *X* (*t*) = *N* = 1, where *X* ∈ {*S, E, X*_*I*_, *R*_*A*_, *R*_*S*_, *D*} labels the population compartments and *X*_*I*_ ∈ {*P, I*_*Sm*_, *I*_*A*_, *I*_*Ss*_, *Y*_*Sm*_, *Y*_*A*_, *Y*_*Ss*_, *H, H*_*ICU*_} labels the subset of compartments that are infectious.

**Table 2:** Disease states included in the model. The dependence on time *t* is implicitly assumed.

| Disease State $X$ | Description | Infectious $X_I$ | Diagnosed |
| --- | --- | --- | --- |
| $S$ | Noninfected and susceptible. | No | No |
| $E$ | Exposed and infected but not yet infectious. | No | No |
| $P$ | Presymptomatic infectious. | Yes | No |
| $I_{Sm}$ | Nondiagnosed Infected with mild symptoms. | Yes | No |
| $I_{Ss}$ | Nondiagnosed infected with severe symptoms. | Yes | No |
| $Y_{Sm}$ | Diagnosed infected with mild symptoms. | Yes | Yes |
| $Y_{Ss}$ | Diagnosed infected with severe symptoms. | Yes | Yes |
| $H$ | Hospitalized not in the intensive-care unit. | Yes | Yes |
| $H_{ICU}$ | Hospitalized in the intensive-care unit. | Yes | Yes |
| $I_A$ | Nondiagnosed infected asymptomatic. | Yes | No |
| $Y_A$ | Diagnosed infected asymptomatic. | Yes | Yes |
| $R_S$ | Recovered who were symptomatic. | No | Yes & No |
| $R_A$ | Recovered who were asymptomatic. | No | Yes & No |
| $D$ | Those who have died. | No | Yes & No |

In the PBM, the susceptible that get infected enter state *E* at a rate known as the force of infection *λ* described in the section A.5. As for all the transition rates in our PBM, *λ* is specified as a per-person transition probability per unit time. Figure 4 illustrates disease progression in the early stages of COVID-19 infection. They progress to the presymptomatic infectious state *P* at a rate *ν*. As an individual-level interpretation of this transition, the mean duration, also known as the mean dwelling or sojourn time that newly infected individuals stay in disease state *E* is given by *ν*^−1^. Those in the presymptomatic infec-tious state *P* either remain asymptomatic and transition to state *I*_*A*_ at a rate *γ*_*A*_, or develop mild symptoms and transition to state *I*_*Sm*_ at a rate *γ*_*S*_. At the individual level, the mean duration that infected individuals stay in disease state *P* is given by [*γ*_*S*_ + *γ*_*A*_]^−1^, and the probability of developing mild symptoms is given by *γ*_*S*_ · [*γ*_*S*_ + *γ*_*A*_]^−1^. We assume that testing the presymptomatic for COVID-19 results in a false negative outcome. Consequently, all those who are presymptomatic are unaware of their infected and infectious state. The formulation of the first three ODEs describing our PBM are

**Figure 4:**
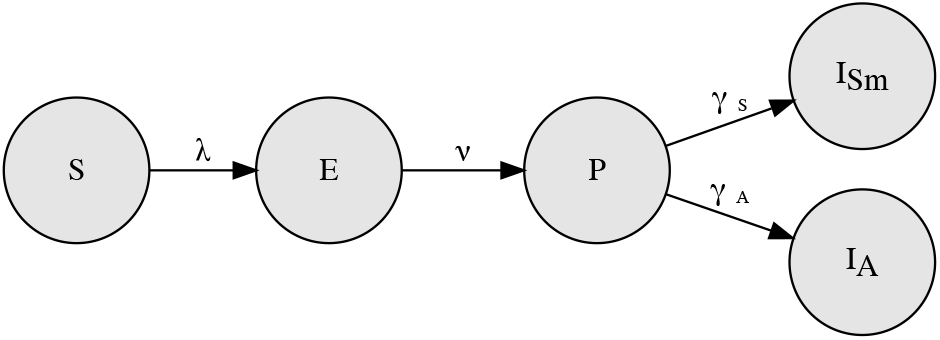
Initial disease progression stages

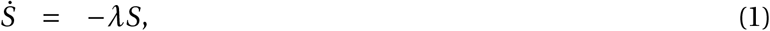

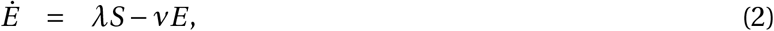

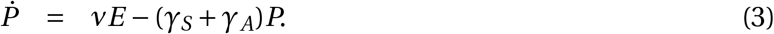

The asymptomatic and unaware of having been infected, *I*_*A*_ progress to the recovered state *R* at rate *ξ*_*A*_. The asymptomatic are assumed to stay infectious until they recover. Figure 5 illustrates the disease progression of the asymptomatic. A proportion *ζ*_*A*_(*t*) · [*ζ*_*A*_(*t*) + *ξ*_*A*_]^−1^ of the asymptomatic are diagnosed with having COVID-19. This proportion of individuals transition to state *Y*_*A*_.

**Figure 5:**
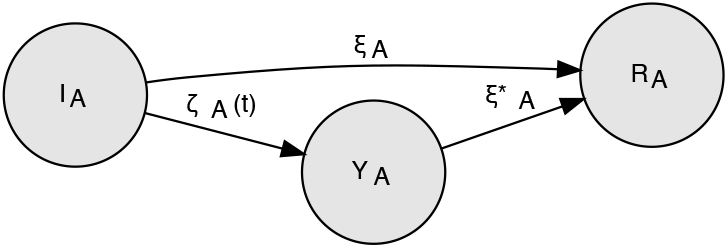

As described in section A.3, the testing rate *ζ*_*A*_(*t*) is not constant but implicitly depends on time, due to dynamic testing policies.

These individuals most likely get tested because they suspect or are informed by health-care workers engaged in contact-tracing to have recently been in contact with someone diagnosed with COVID-19. Those who enter disease stage *Y*_*A*_ progress to the recovered state *R* at rate 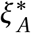. The transition rate 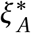 is faster than *ξ*_*A*_ and accounts for the elapsed duration of mild symptoms prior to being diagnosed. These diagnosed asymptomatic are aware of having been infected and of being infectious. Therefore, they are assumed to engage in increased social distancing behavior described in the section A.3 on the disease transmission model. The ODEs describing the asymptomatic disease progression are

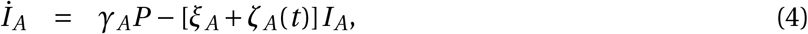

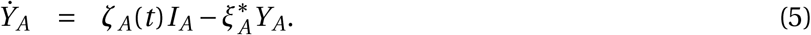

Figure 6 illustrates the disease progression of the symptomatic. The non-diagnosed mildly symptomatic, *I*_*Sm*_ either progress to the recovered state *R* at a rate *ξ*_*m*_ or develop severe symptoms and progress to state *I*_*Ss*_ at a rate *ν*. Some are tested at a rate *ζ*_*S*_(*t*) and diagnosed with having COVID-19. These transition to state *Y*_*Sm*_ and become aware of their infection state. As before, *ζ*_*S*_(*t*) is not a constant but is assumed to implicitly depends on time. Our model assumes that those mildly symp-tomatic that get diagnosed will also progress to the recovered state *R* at a rate 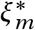 or develop severe symptoms and transition to state *Y*_*Ss*_ at rates *ν*^*^. These rates are respectively faster than *ξ* and *ν* to account for the elapsed duration of mild symptoms prior to being diagnosed. The ODEs describing the non-hospitalized with mild symptoms are

**Figure 6:**
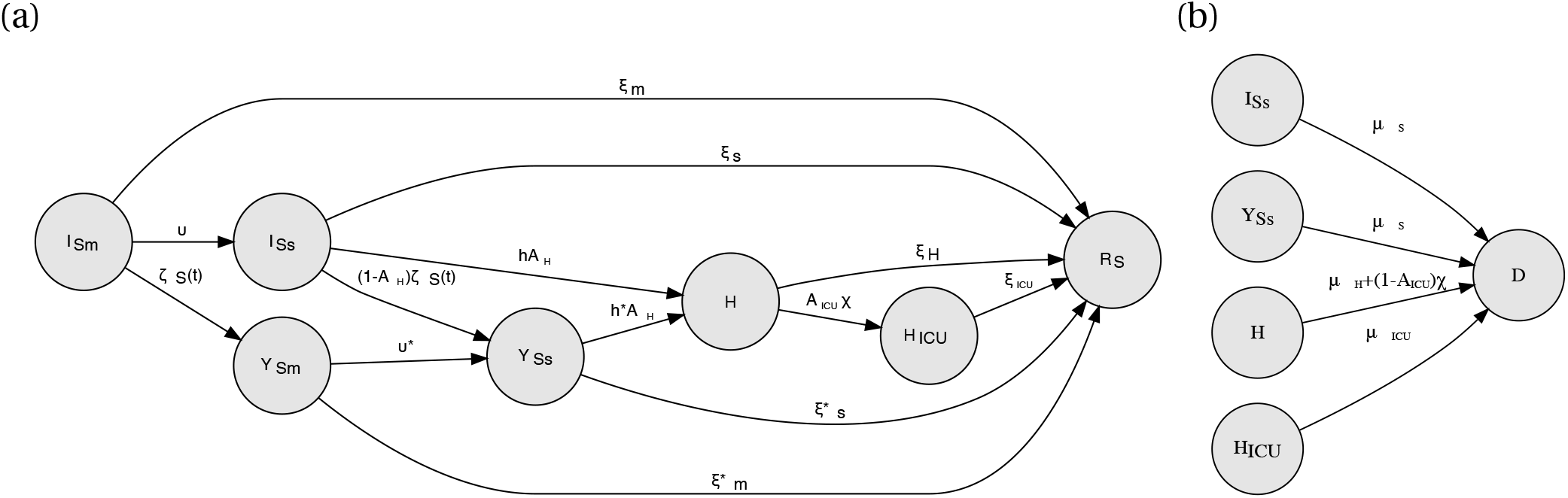
Model flow of disease progression of the infected symptomatic with transitions that lead to the recovered state *R* and the died state *D* shown respectively in panels a and b.

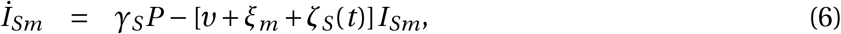

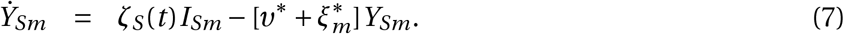

Most of the non-diagnosed severely symptomatic *I*_*Ss*_ are hospitalized at a rate *hA*_*H*_. When they enter the hospital they are diagnosed with COVID-19. The percentage of severe cases hospitalized remains constant as long as the hospital has not reached its capacity in terms of available beds. The dichotomous variable *A*_*H*_ indicates whether the hospital is accessible to COVID-19 patients (*A*_*H*_ = 1) or whether it has reached its bed capacity and no longer accepts new COVID-19 patients (*A*_*H*_ = 0). If the hospital is at capacity, the severely symptomatic patients that would have otherwise been hospitalized are tested at a rate (1 − *A*_*H*_)*ζ*_*S*_(*t*). Once the hospital is no longer at capacity, these diagnosed patients are hospitalized at a rate *h*^*^ *A* which is faster than rate *h* as these diagnosed patients are likely to have waited longer to be hospitalized than the non-diagnosed. A minority of the severely symptomatic never access the hospital. Some of these patients recover but most die at home. We assume that the transition rates to the recovered state *R* is *ξ*_*s*_ and to death *D* is *µ*_*s*_. These rates also apply to those diagnosed and severely symptomatic as the majority of those in state *Y*_*Ss*_ are diagnosed while having mild symptoms. The ODEs describing the non-hospitalized with severe symptoms are

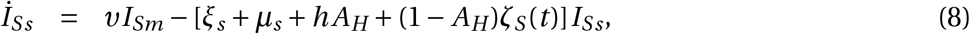

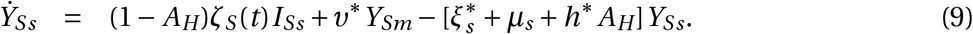

Patients that are hospitalized may develop critical symptoms. Like for the hospital, the ICU can also reach capacity, at which point it takes no more patients until it has free beds. Provided there are available ICU beds, these hospitalized critical patients transition into the ICU at a rate *𝒳A*_*ICU*_. As with hospital accessibility, we use the dichotomous variable *A*_*ICU*_ to indicate whether the ICU is accessible (*A*_*ICU*_ = 1) to COVID-19 critical patients or whether it has reached capacity (*A*_*ICU*_ = 0). Hospitalized patients that do not require the ICU either transition to the recovered state *R* at a rate *ξ*_*H*_, or transition to death *D* at a rate *µ*_*H*_. The transition rate *µ*_*H*_ is assumed to be small because very few patients die in the hospital without having accessed the ICU first, provided that the ICU is not at capacity. We assume that patients who recover in the ICU move immediately to the recovered compartment rather than back to the hospital. This ensures that individuals do not make multiple trips to the ICU. The transition rate to the recovered state *R* is *ξ*_*ICU*_. Therefore, the time spent in the *H*_*ICU*_ compartment represents both the ICU and the time spent recovering after intensive care in the hospital. The actual transition rates from the hospitalized to the recovered state *R* and death *D* depend on whether the ICU is accessible. When the ICU is closed, we assume that all those who required ICU access will die until the ICU is reopened. Hence, the general transition rate from *H*_*ICU*_ to death *D* is *µ*_*H*_ +*𝒳*(1− *A*_*ICU*_). The ODEs describing the hospitalized are

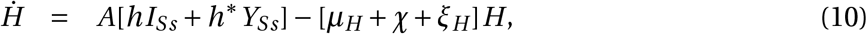

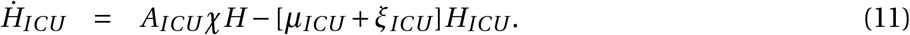

When the hospital is at capacity (*A*_*H*_ = 0), patients that develop critical symptoms are more likely to die at home. This requires modifying the rate *µ*_*s*_ based on the hospital accessibility indicator variable *A*_*H*_. The implementation of our PBM can take this into account where the death rate *µ*_*s*_ in state *I*_*Ss*_ is increased to *µ*_*s*_ + *h𝒳*/(*𝒳* + *ξ*_*H*_). A similar increase in the death rate applies in state *Y*_*Ss*_. However, unlike for the ICU, the duration of having no accessibility to the hospital is likely to be short because hospitals can adapt spaces and create new bed accommodations in a way that is not possible for the ICU. Hence, the ODEs we present and describe here do not consider this increased mortality rate. Therefore, the ODEs describing the recovered and those that die are

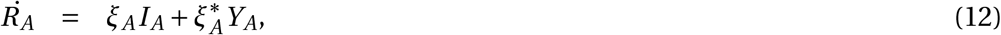

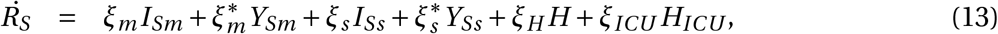

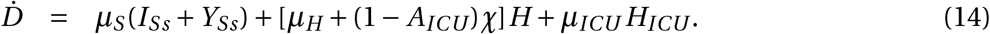

We assume that the expected time for the diagnosed and asymptomatic *Y*_*A*_ to transition to the recovered state *R* is shorter than the expected time for the same transition to the recovered state of the undiagnosed asymptomatic *I*_*A*_. Similarly, the expected time for *Y*_*Sm*_ to transition to either state *Y*_*Ss*_ or the recovered state *R* is shorter than the expected time for those in *I*_*Sm*_ to transition to either state *I*_*Ss*_ or the *R* is. We model this by setting 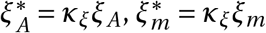 and *ν*^*^ = *κ*_*ξ*_*ν* where the value of *κ*_*ξ*_ is sampled in a range of values greater than one. As for interpretation, when (*κ*_*ξ*_−1)/*κ*_*ξ*_ is multiplied by the expected duration before the next clinical disease progression, it gives the expected time duration when an infected person is likely to be diagnosed with COVID-19.

It is important to note that we have used a single compartment to represent each disease state. Since our model tracks populations’ flow between compartments rather than individuals, implicitly, the sojourn or dwelling time distribution for each compartment is exponential. Hence, it is a Markov process with the following individual-level interpretation. The sojourn times and the transition rate for the next disease progression do not depend on the history or path taken in how an individual reached a specific disease state, nor does it depend on the elapsed time they have spent in each disease state. This property can have profound consequences in the disease progression dynamics, ultimately affecting transmission. The boxcar method can alleviate the strong assumption of exponential dwelling time distributions substituting the exponential distribution with an Erlang distribution, which is more pathologically realistic [12]. This method requires breaking each disease state into a series of concatenated compartments of the same disease state and shorter internals. By making the shorter intervals equal in duration we obtain an Erlang dwelling time distribution. Conversely, the shorter internals could have different duration to approximate other types of dwelling time distributions. For example, a Weibull distribution can be approximated by a series of concatenated compartments with decreasing sojourn times [13]. A model that uses the box-car method retains its Markov property and has more realism in terms of progression rates. However, using this method the model becomes computationally expensive and has diminishing benefits for disease models like our PBM, which has many disease compartments, each with relatively short dwelling times.

### A.2 Loss of immunity

The description of our model so far follows the general framework of an SEIR model. However, research indicates that immunity to COVID-19 is not permanent nor enduring and can last less than a year. To simulate this, we adopt an SEIRS model, whereby those who recover can become susceptible again. Since the rate of loss of immunity is slow with a time scale of months or years compared to the disease progression rates with time scales of days, having those who have recovered transition directly to the susceptible state is too rudimentary. This is because, as mentioned in the previous section, progression rates from one compartment to the next assume an exponential dwelling time distribution, and this assumption for the dwelling time of being recovered and immune is highly inaccurate. Hence we adopt a simple first-order boxcar correction whereby we include an intermediate or buffer recovered compartment *R*_*B*_, which recovered people transition into before losing immunity and returning the susceptible population pool *S*. Hence, equation 1, 12 and 13 are modified, and we have an ODE describing the dynamics of the *R*_*B*_ compartment

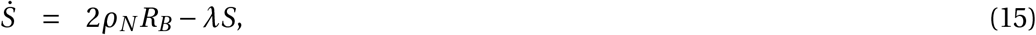

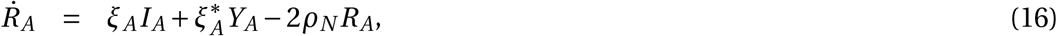

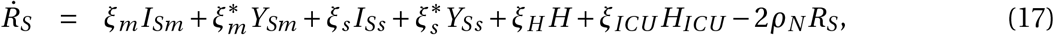

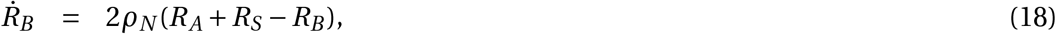

where *ρ*_*N*_ is the rate of loss of naturally acquired immunity. Extending the first order boxcar method to higher orders can accomplish more realistic distributions of natural immunity duration. It would require increasing the number of intermediate buffers *R*_*B*_ and correspondingly decrease the transition multiplicative factor from 2 to *n* + 1 where here *n* represents the number of intermediate buffers.

In our model, those who lose immunity return to the susceptible state and are treated as any other susceptible. This means that our model does not consider a better prognosis due to residual immunity from the previous infection for people infected a second time. Hence, this effect leads to more pessimistic long-term and multiple-year model dynamics.

### A.3 Testing

Our model assumes a daily per-person rate *ζ*(*t*) of being tested for COVID-19 on the population. People in different disease states have different demands for getting tested. For example, we assume that people that are aware of having had COVID-19 and have recovered do not seek a test. We assume that people tested in the exposed and presymptomatic infectious state are not diagnosed with COVID-19 as their test results in a false negative outcome. The specific rates *ζ*_*S*_(*t*) and *ζ*_*A*_(*t*) respectively represent the daily per-person detection and diagnosis rates for the symptomatic and asymptomatic populations. Those with mild and severe symptoms are more likely to get tested than those who are asymptomatic. Hence *ζ*_*S*_(*t*) > *ζ*_*A*_(*t*). The rate *ζ*_*S*_(*t*) only applies to those not yet hospitalized who have either mild or severe symptoms. We assume that those who are hospitalized get tested if they did not previously test positive. The rate at which the asymptomatic population seeks a test can be considered the same for the susceptible population. However, due to testing-and-tracing efforts, it is instead likely to be marginally larger, and hence *ζ*_*A*_(*t*) > *ζ*(*t*). Under the assumption of unconstrained testing rates, these parameters are treated as constant and depend on the base-case demand for testing. This means that anyone who seeks a test is tested. Therefore, we assume testing capacity can accommodate the growth of the epidemic, including the exponential phase.

Our PBM can model both the initial and later stages of the pandemic. The assumption of unconstrained testing rates is best suited to model the later stages of the epidemic. The results presented in this paper consider unconstrained testing rates. However, for completeness, we proceed to describe the capability of our PBM in modeling the early phase of the pandemic where the number of available testing kits was limited. The implementation of our PBM can consider settings with constraints in the daily number of available testing kits. This number is assumed to start low and grow linearly to a predefined maximum daily testing rate capacity. Thus, under these settings, the actual rates describing the testing and diagnosis rates from each disease state are reduced based on the capacity constraint. We assume that people who have been hospitalized take priority and are tested first. The remaining number of testing kits is then used to test those with severe symptoms. After that, the remaining number of testing kits is used to test those with mild symptoms, followed by the asymptomatic. This approach requires specifying a constant proportion of testing kits used to successfully identify COVID-19–positive compared with those that are COVID-19–negative.

### A.4 Additional Outputs

Our PBM extracts and combines incidence rates that flow into different compartments to track output cumulative quantities by population strata that do not affect the dynamics. These outputs include

1. True cumulative case counts:

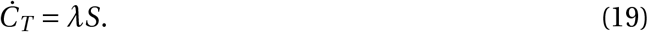
2. Reported cumulative case counts:

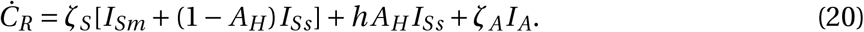
3. Cumulative number of people tested:

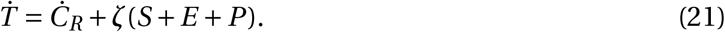
4. Reported recovered:

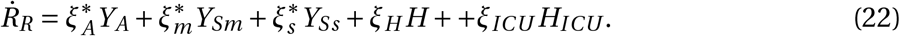

We assume that all diagnosed COVID-19 cases that do not result in death are reported as recovered cases.
5. Reported deaths:

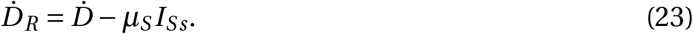

We assume that all diagnosed COVID-19 cases that result in death are reported. All suspect deaths due to COVID-19 of those with severe symptoms that did not get diagnosed and did not progress to the hospital are assumed to be unreported.
6. Reported Case Fatality Rate (CFR):

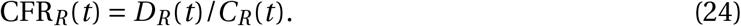
7. The Infection Fatality Rate:

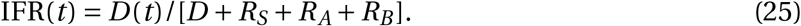

### A.5 Modeling SARS-CoV-2 transmission

Following the approach taken in standard compartmental models of infectious diseases, the force of infection *λ* describes SARS-CoV-2 transmission. The force of infection is characterized by how infectious people in each disease state infect others. In our formulation, the force of infection is the product of two parameters. The first parameter is the contact mixing rate, representing the number of daily contacts people make with others. The second parameter is biological transmissibility, which defines the probability of transmission between an infectious and a susceptible person when they come in contact. Each disease state would have a different content mixing rate and transmissibility. We express the force of infection as

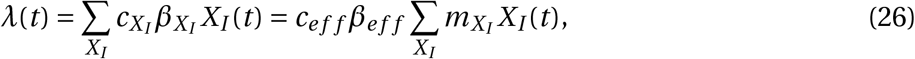

where the coefficient 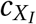 represents the social mixing contact rate and 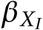 represents the transmissibility of infectious people in disease state *X*_*I*_.

The expression for the force of infection simplifies by assuming an effective contact rate *c*_*eff*_ and an effective *β*_*eff*_ transmissibility. The product of these two *c*_*eff*_ *β*_*eff*_ is assumed to characterize the rate of infections caused by an undiagnosed asymptomatic infected person in either state *P* or *I*_*A*_. Rates of infections in the other disease states are characterized using 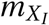 coefficients that give the multiplicative effect on infectivity with respect to the primary infectious state or an asymptomatic state. For example, *m*_*Ss*_ gives the overall average multiplicative infectivity of a symptomatic severe individual relative to an asymptomatic individual. This multiplicative factor combines the effect of decreased social mixing with increased biological transmissibility. We choose the asymptomatic untested individual as our reference because they are unaware of their positive status and thus do not change their social mixing behavior acting as though they are not infected. Hence 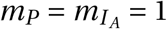. We estimate the values of the multiplicative factor of the other disease stages by considering how the transmissibility and the social mixing contact rate change relative to the presymptomatic case.

Changes in viral load are used to estimate the transmissibility of each of the disease states. Studies have shown that viral loads peak in the primary infectious stage and decrease monotonically after the onset of symptoms. Hence, we assume the inequalities 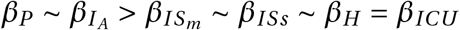. In terms of mixing rates we assume that 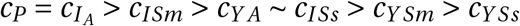. See section B.2.3 for more details on relative infectivity. Those who are in the presymptomatic or asymptomatic state are unaware of their positive status and thus act as though they are not infected. However, those who have symptoms will reduce their contacts as their conditions become more severe or receive a positive test result. Likewise, tested asymptomatic people also will begin to practice increased social distancing to protect their social contacts.

Under ideal conditions, the physical contact rate that leads to disease transmission between health care workers and COVID-19 patients *c*_*H*_ and *c*_*ICU*_ would be close to zero. However, given the shortage of personal protective equipment during the early stages of the epidemic, contact safety precautions in health care settings may not have always been perfectly adhered to. We consider a range of assumptions for *c*_*H*_ and *c*_*ICU*_.

The value of the effective infectivity *c*_*eff*_ *β*_*eff*_ and its range is estimated from the basic reproductive number *R*_0_. This number is defined at the individual-level and represents the average number of secondary infections caused by an infectious person during the disease invasion phase. This phase represents the early stage of the epidemic when susceptible individuals surround each infectious person. At the population-level, *R*_0_ represents a threshold parameter. Whether its value is larger or smaller than one, this threshold parameter indicates whether an outbreak will invade the population and become an epidemic or whether it is likely to extinguish before becoming a full-blown epidemic. The distinction between the individual-level definition and the basic reproductive number’s thresh-old property does not always align and can be very consequential, leading to incorrect conclusions. A large body of literature provides a detailed discussion on the basic reproductive number, its uses, limitations, and misconceptions [14–18].

In simple mathematical models of infectious diseases, such as Susceptible-Infected-Removed (SIR) or SEIR models, *R*_0_ can be expressed as the product of three terms *R*_0_ = *cβτ*_*I*_ where *τ*_*I*_ represents the duration of the infectious phase. This expression assumes that the contact rate *c* and the transmissibility *β* takes the same value for all the infectious compartments. This is the case for the SIR and SEIR models but it is not the case for our COVID-19 PBM. Our COVID-19 model considers more-infectious compartments with different contact mixing and transmissibility values and compartments that branch off from each other. We express *R*_0_ in terms of *c*_*eff*_ *β*_*eff*_ as

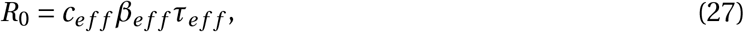

where *τ*_*eff*_ represents a typical time-scale that considers the diversity of both disease transmission and progression across the different disease states. It can be interpreted as the effective infectious period for an equivalent SEIR model with a single infectious compartment, with contact rate *c*_*eff*_ and the transmissibility *β*_*eff*_. Consequently, in our model *τ*_*eff*_ is not equal to the duration of the infectious period *τ*_*I*_ because the former accounts for the changes in transmissibilities in each infectious compartment, namely the *m* multiplicative factors. The expression for *τ*_*eff*_ is given at disease invasion and hence considers the case where testing rates *ζ*_*A*_(*t*) and *ζ*_*S*_(*t*) are zero. The next section, appendix A.6, describes the next-generation method and gives the mathematical expression for *τ*_*eff*_ at disease invasion in terms of the transmission and progression parameters. Hence, by knowing the input transmission and progression parameter values, we can compute the duration *τ*_*eff*_. By knowing the value of this duration and an estimated input value of *R*_0_ we can compute the value of *c*_*eff*_ *β*_*eff*_.

To extract an estimate for the value of *R*_0_, we use the number of reported cases during the epidemic’s early stages. In the disease invasion phase, the growth in cases is exponential. Hence, the log of the case counts and the log of the death counts increase linearly with time. An estimate for the growth rate *r* is obtained by linear regression of the log of these counts with time. Mathematically, *R*_0_ is related to *r* by the following expression

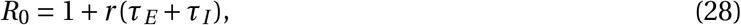

where *τ*_*E*_ + *τ*_*I*_ represents the typical duration for which a person is infected and is the sum of the duration of the noninfectious incubation phase *τ*_*E*_ and the duration of the infectious phase *τ*_*I*_ [17]. The value of *τ*_*I*_ depends on the disease progression times and, more specifically, on the dwelling or sojourn times of compartments representing infectious states.

As disease dynamics progress beyond the disease-invasion phase and natural immunity develops, we use a time-dependent parameter known as the effective reproduction number *R*_*t*_. Tracking the effective reproductive number dynamics is informative because it exhibits the same threshold properties as the basic reproductive number. Hence, if its value is above 1, the disease is spreading and is not under control. At the individual-level *R*_*t*_, it represents the average number of new infections caused by a single infected individual at time *t* in the partially susceptible population. Generally, it is found by multiplying *R*_0_ by the proportion of the population that is susceptible. For our model this means substituting equation 30 for *τ*_*eff*_ into equation 27 for *R*_0_ and multiplying by *S*(*t*). However, equation 30 provides an approximate value for *τ*_*eff*_ which is only exact at disease invasion. This is because *R*_0_ is defined at disease invasion, and this allows us to remove unnecessary details from the calculation, which do not matter at disease invasion. For example, we assume that no-one is being tested at disease invasion and that the hospital is at capacity. Hence, complexities in behavioral changes due to testing and changes in the dynamics due to hospital capacity constraints can be neglected. However, when computing *R*_*t*_ these details become important and dynamically change the typical time-scale *τ*_*eff*_ of the disease. We can obtain an equation for *τ*_*eff*_ that replaces equation 30 which is more generally valid, and our model can rely on it. However, it is a very lengthy algebraic expression. We discuss this further in appendix A.6.

The estimation of the value of the growth rate *r* and hence of *R*_0_ using a linear regression approach has limitations. During the disease invasion exponential growth phase, case reports are not very reliable because of backlogs and limited testing capacity. By the time testing rates and capacity have stabilized, many jurisdictions were already in a stage where they had implemented social distancing. A better choice is to use the death count data. However, when the linear regression approach is used to find the *R*_0_ for the different U.S. states we have found that some states produced an unacceptably bad fit to the data. For these states, we assumed that the value of *R*_0_ is between two and four based on the population density of the state. For these cases, the value of *R*_0_ is assumed to range from two for the state with the lowest population density to four for the state with the highest population density.

### A.6 The expression for *τ*_*eff*_

To find the expression for *τ*_*eff*_, we apply the next-generation method [19, 20] to the model described by the set of coupled ODEs given in Eqs 1-14. We first rearrange the order of our ODEs and focus only on the equations describing infection states. We then construct the matrices **M** and **V** respectively, describing the disease transmission and progression terms. The equations, together with the matrices, are shown on the next page. A representation of the Jacobian matrix **J**^*^ of our system of ODEs taken at disease-free equilibrium point (i.e., *S* = 1) is given by

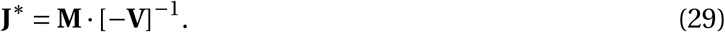

The expression for *R*_0_ is found by finding the largest eigenvalue of **J**^*^. At the disease-free equilibrium point, and soon after, at disease-invasion, the hospital and ICU is not at capacity and there is no testing for COVID-19. Hence the expression for *R*_0_ is found by setting *A*_*H*_ = *A*_*ICU*_ = 1 and *ζ*_*S*_ = *ζ*_*A*_ = 0). Using this simplification we find that

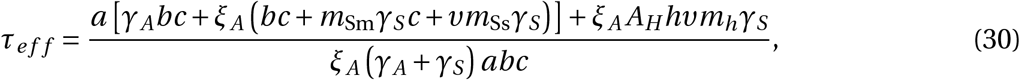

where the coefficients *a, b* and *c* are given by

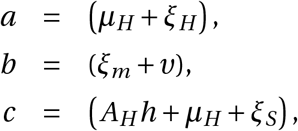

and

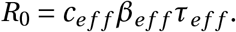

We can also find an expression for the effective reproductive number *R*_*t*_. Generally *R*_*t*_ = *R*_0_*S*. However, we cannot use equation 30 for *τ*_*eff*_ and to get *R*_*t*_ because it is only valid at disease invasion where we assumed no testing and full accessibility to the hospital and ICU. We can derive a complete expression for *τ*_*eff*_ that considers testing rates and hospital accessibility using the next-generation method. Such expression is algebraically long and complicated, and we choose not to include it here. A Mathematica notebook providing the full expression is available upon request.

The ODEs of our model can be reordered as follows:

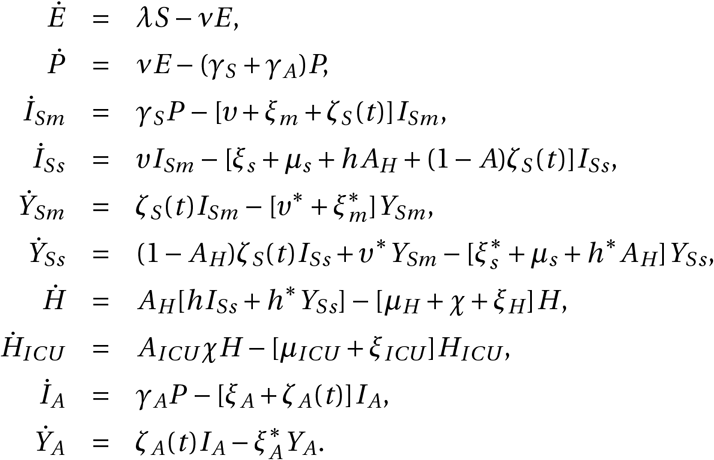

Using this order, the matrix describing the transmission terms **M** is given by

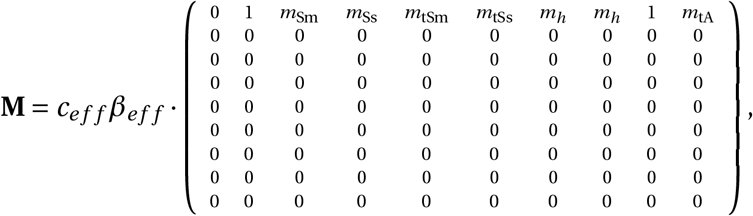

and the matrix **V** describing the progression terms is given by

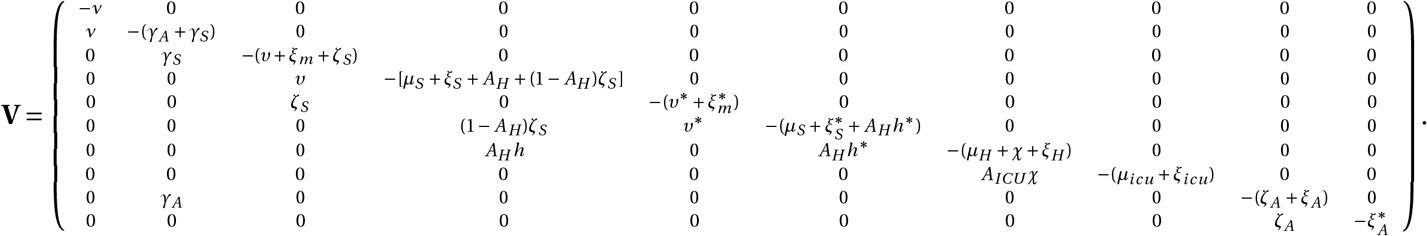

We note that although *τ*_*eff*_ expresses a time scale, it includes the multiplicative coefficients *m*. Thus, *τ*_*eff*_ gives the time scale of the whole infectious state’s Q% when all multiplicative coefficients *m* are set equal to one. Using a more direct approach instead of the next-generation matrix approach, we verified that the expression for Q-.. does indeed give Q% when all multiplicative coefficients D are set equal to one. Because

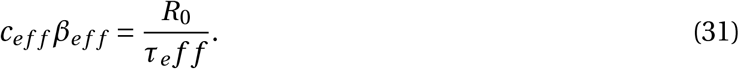

### A.7 The Population Stratified Model

The population in our model can be partitioned into subpopulations. We consider multiple population groups or strata within each compartment. Each stratum specifies the population-based on common characteristics, such as demographic, social, economic, and pathological states. The specific strata we consider are described in section B.1. The structure of the population stratified model is expressed as an array of ODEs, where the disease progression dynamics for each stratum are expressed by equations 1-14. The reformulation to a strata-dependent PBM extends the model from the more conventional version of a single-strata compartment model that assumes homogeneous mixing and implicit interactions within the population.

The modeled population is described by the proportion of people in each stratum. These proportions are given by the one-dimensional array ***v*** where the element *v*_*i*_ represents the proportion of the population belonging to strata *i*. Since we are tracking population densities, the sum of all the elements of the ***v*** is equal to one. We will refer to ***v*** and other strata-specific one-dimensional arrays as vectors in the disease state space *X*. Therefore, each disease state is also expressed as a vector. For example, the vector ***P*** (*t*) represents the presymptomatic state, and the element *P*_*i*_ represents the proportion of the presymptomatic population belonging to strata *i*.

Some parameter values and settings that enter the model differ across the strata. The heterogeneity introduced by the strata-dependent parameters plays a crucial role in the disease progression and transmission dynamics and is used to better characterize the epidemiology of COVID-19. Differences across strata include parameter values describing pathological transition rates, including the proportion of the infectious in each stratum that stay asymptomatic (i.e., *γ*_*A*_ and *γ*_*S*_), the proportion of the symptomatic that develop severe symptoms and need hospitalization (i.e., *ν*), and the fatality rates (i.e., the *µ* parameter values). The flexibility of our model allows users to easily specify other parameters that depend on the strata. For example, protective behaviors such as willingness to get tested can be specified by stratum.

The strata-dependent parameters are also expressed as vectors and are used to specify the strata-specific transition rates between disease states. For example, the vector giving the strata-specific transition rates from state ***P*** to state ***I***_*A*_ is expressed as *γ*_*A*_ ⊙ ***P***, where ⊙ denotes the element-wise multiplication. Equivalently, this can be expressed by matrix multiplication as diag(*γ*_*A*_) · ***P*** where the operator diag represents vector diagonalization. By following this notation, the PBM’s ODEs can be expressed in vector notation, remaining mathematically concise.

Heterogeneity in disease transmission is introduced by the strata-dependent mixing contact rates describing the variations in how people belonging to the different population strata mix with each other. This heterogeneity is described by a mixing matrix ***M***. The matrix element *M*_*i j*_ denotes the proportion of contacts of individuals in population strata (or row) *i* with those in population strata (or column) *j*. The sum across each row in the mixing matrix is a sum of proportions, and hence equals one (i.e., Σ_*j*_ *M*_*i j*_ = 1). In addition to the mixing matrix, we have a normalized contact vector ***κ***. The vector element *κ*_*i*_ denotes the proportion of all daily contacts (or duration of contacts) in the population made by individuals in stratum *i*, and hence, the sum of the vector elements is one (i.e., Σ_*i*_ *κ*_*i*_ = 1). The matrix multiplication of the diagonalized vector ***κ*** with ***M*** gives the contact matrix ***K***, expressed by

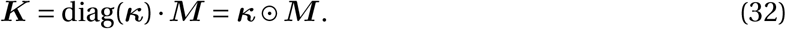

The contact matrix ***K*** is a symmetric matrix where the sum across the rows is equal to the vector ***κ***. For off-diagonal elements *i* and *j*, the sum *K*_*i j*_ +*K*_*ji*_ gives the proportion of all daily contacts between strata *i* and *j*. For diagonal elements, the same proportion is given by *K*_*ii*_. Under the disease-free status-quo conditions everyone is susceptible. Hence ***v*** = ***S***, and the overall daily effective contact rate is propositional to ***S***^T^·***K*** ·***S***. During the disease transmission dynamics, we are instead interested in the contacts between the susceptible population ***S*** and each of the infectious states ***X***_***I***_. Hence the daily effective contact rate between susceptible and infectious people is proportional to 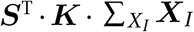. We denote the coefficient of proportionality by *k*_*λ*_. Taken together with transmissibility *β*_*eff*_ and the transmissibility multiplier 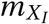, we can express the force of infection vector for our strata dependent PBM as

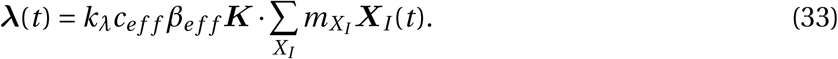

Equation 33 replaces equation 26 for our strata-specific PBM and the transition rate from *S* to *E* given in equation 1 is re-expressed as

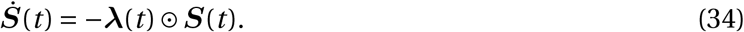

The coefficient of proportionality *k*_*λ*_ is related to the base-line total daily contact rates. We estimate *k*_*λ*_ at disease-invasion by calibration to the observed data and assume that it stays constant during the dynamics. Instead, changes in mixing rates due to social-distancing NPI and behavioral responses are accounted for by changes in the contact matrix ***K***.

### A.8 Mixing Modes

People mix in different settings, or mixing modes. Modes have different levels of social interaction and different strata compositions. For instance, in schools mixing is primarily between children, whereas in commercial settings mixing occurs between all ages. Our PBM considers six different modes of social mixing: household, school, work, commerce, leisure, and other. Based on the data described in Section B.1 we can create matrices describing the average daily contacts between each stratum in each mixing mode. We decompose these matrices into a set of row normalized mixing matrices ***M***_*m*_, column normalized contact vectors ***κ***_*m*_, and scalar mode weight *w*_*m*_ for each mixing mode labeled by the index *m*. The total contact matrix, *K* is a weighted sum of the mode-specific contact matrices ***K***_*m*_, and given by

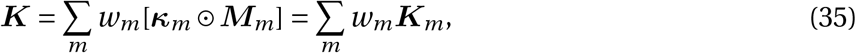

The weights, *w*_*m*_ give the proportion of contacts (or duration of contacts) of how people mix over the different mixing modes. Under the disease-free status-quo conditions these weights sum to one, hence Σ_*m*_ *w*_*m*_ = 1.

### A.9 Modeling Nonpharmaceutical interventions (NPIs)

When the initial outbreak becomes an epidemic, governments may choose to impose nonpharmaceutical interventions (NPIs) which limit how people can mix. The goal of NPIs is to delay and reduce the peak number of cases and hospitalizations per day by shifting individuals to locations where there are few unique contacts. Hence, we model the effects of NPIs by decreasing the weights associated with various mixing modes. For example, an NPI policy that closed schools, restaurants, and bars is modeled by reducing the weights associated with the school, commerce, and leisure mixing modes. Mixing matrices measure the number of unique contacts, not time spent in a mixing mode, therefore weights are not conserved. NPI interventions, such as confining people to their households, reduce the total number of unique contacts such that the mixing mode weights sum to less than one. To model the impact of reduced mixing from NPI level *n* on mode *m*, we define a diagonal matrix 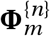. The diagonal elements of 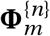 specify the reduction in mixing for each stratum in mode *m* relative to the disease-free state. For interventions that apply to all strata (i.e. where each stratum changes their mixing by the same proportion), such as the closure of schools, all diagonal elements of 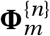 have the same value. In cases where interventions apply to some strata and not others: such as when only front-line essential workers are expected to attend their workplaces, the diagonal elements of 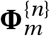 take on different values, each specifying the strata-mode specific impact of the NPI. Hence the expression for ***K***^{*n*}^ that accounts for the impact of NPIs is:

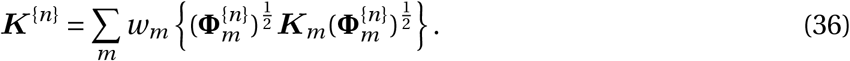

In the disease-free state, specified by NPI-level one (i.e., *n* = 1) the 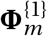 matrix is equal to the identity matrix where all diagonal elements are equal to one.

The matrix 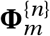 is square-rooted and then multiplied on either side of the contact matrix ***K***_*m*_ so that the contact matrix is symmetric. The matrix 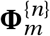 represents multiplicative factors that reduce the strength of the interactions along the network edges. It is obtained by the individual-level reductions specified on the network vertices. Two vertices bound each edge, and hence the overall edge-level reduction of the interactions is given by multiplying the two vertex-level reductions. These reductions are represented by the term 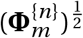 in mixing on either side of the strength of the edge, and given by the matrix ***K***_*m*_.

We allow the effectiveness of NPIs to vary by state. The effectiveness of the NPIs, *θ*, is defined through a calibration process. We narrow the ranges used as priors in the calibration process by exploring a wide range of parameter combinations using a Latin Hypercube Sample and by observing that values outside those ranges consistently produced biased death results. The calibrated contact matrix 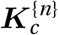 is:

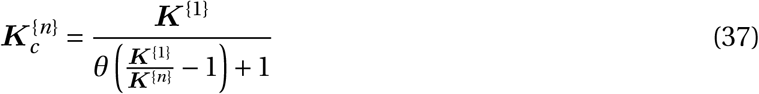

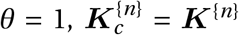. For other values, *θ* approximately geometrically scales the distance between ***K***^{*n*}^ and zero mixing, while leaving 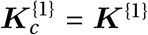 for any *θ*. For instance, a value of *θ* = 2 yields a 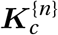 approximately half the size of ***K***^{*n*}^ for *n* ≠ 1. This functional form allows us to account for the physical and cultural differences between states which may yield different NPI effectiveness and compliance levels.

After the initial wave of lockdowns in March and April 2020, many states slackened restrictions. However many behaviors to reduce transmission, including mask-wearing, aversion to crowded spaces, and other adaptation measures remained after the lockdowns were lifted. To account for this we compute the final contact matrix 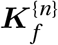 as a weighted average of the contact matrix under the highest NPI 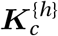 and the current NPI 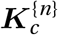

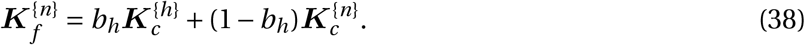

The relative weight *b*_*h*_ of the highest NPI matrix is calibrated through the same process as the NPI effectiveness parameter. Finally, to prevent NPIs from changing instantaneously in the model and causing discontinuities inappropriate for an ODE, we make the NPI level *n*_*c*_ a continuous stock variable with rate 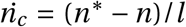, where *n*^*^ is a target NPI level and *l* determines how fast the NPI level can be changed. Therefore, the mixing matrix we use is a weighted average between the ceiling NPI level mixing matrix 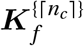 and the floor NPI level 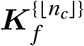 weighted by the distance between *n*_*c*_ and its ceiling.

### A.10 Modeling NPI-induced changes in household mixing

The NPI-specific contact matrices ***K***^{*n*}^ specify average mixing levels across everyone in the population. For some modes this is a reasonable approximation - the sample of individuals in a grocery store is approximately random and is likely different for each visit. However, this is not a good approximation of households, in which the same set of individuals mixes every day. Consider a stringent intervention that confined everyone to their home: the mixing weights for each other mode would drop to zero, but the mixing weights for the household would remain at one (because the number of unique household contacts remains constant). This formulation would model the situation in which all individuals are confined to a single (gigantic) household. To approximate the reality that people are confined to different households, we assume that the number of infections transmitted within the household is proportional to the amount of mixing outside the household. That is:

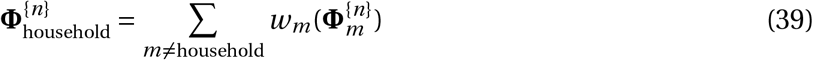

### A.11 Seasonality

The PBM considers seasonality in the transmissibility and social mixing. This is modeled by multiplying *c*_*eff*_ *β*_*eff*_ by a time-varying term denoted by *ϑ*(*t*) that has an average value equal to one over a year. We use a simple sinusoidal function to describe *ϑ*(*t*) given by

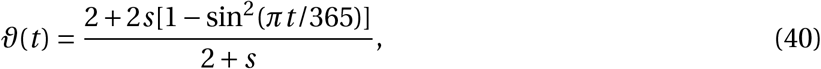

where *s* is a tunable constant. For *s* = 0 we get no seasonal effects and the multiplicative parameter *ϑ* = 1. As *s* → ∞, the multiplicative parameter *ϑ*(*t*) will tend to vary between 0 in early July and 2 in early January with an average value of 1. The sinusoidal function assumes that the seasonal decrease in transmissibility after January mirrors its seasonal increase after July. In reality, changes in transmissibility over the year are not perfectly symmetric as given by a sinusoidal function. Future versions of our model will relax this assumption.

In addition to the smooth change in seasonal transmissibility described by equation 40, our model allows users to specify key date such as national holidays and sports events when people are likely to relax social distancing measures, gather with family and friends, or in large groups, and generally be less compliant to the NPIs.

### A.12 Numerical Integration

The coupled ODEs defining our PBM are integrated numerically to track the dynamics of the population in each compartment as they change over time. Since the model includes many compartments and a wide range of values defining the rates across the compartments, the numerical integration of the ODEs is a stiff problem [8], whereby the numerical solution has its step size limited more severely by the stability of the numerical technique than by the accuracy of the technique. We implemented in R using the deSolve package [9]. DeSolve solves initial value problems for stiff ODEs using FORTRAN solvers of the Livermore family. There are various types of solvers that can be used [6, 7]. We tested the lsoda, vode, and the fourth-order Runge-Kutta method and found that the lsoda method was the most reliable for our purpose.

## B Informing the model

### B.1 Mixing Matrices and Population Strata

We model mixing in the population using mixing matrices. A mixing matrix describes the amount of contact that occurs between each of the population strata. We consider six different mixing locations: household, work, school, commercial, recreational, and other. We use two sources for mixing matrices within the baseline scenario. The first source is a mixing matrix based on the locations of a synthetic population in the city of Portland, Oregon provided by the Network Dynamics and Simulation Science Laboratory (NDSSL) at Virginia Polytechnic Institute and State University [21]. The second data source is based on self-reported survey data. Over the course of one day in eight European countries, 7,290 participants reported 97,074 unique contacts [22]. These results were then extrapolated to create mixing matrices for 152 countries, including the United States [23]. We use the US matrix as our second data source.

These sources represent two different methodologies for quantifying social mixing matrices, and both have limitations. The NDSSL data define a contact as two simulated individuals inhabiting the same sublocation at the same time. This method likely over-weights locations where there are many individuals in low mixing environments, such as workplaces. Further, the data are synthetic and represent Portland on a weekday, so might not be representative of the entire United States. The Prem et al. contacts are self-tracked, so they may over-weight contacts in close environments that are more easily remembered, such as the home. Although they are ostensibly representative of the United States, the original data were from European countries. Additionally, the Prem et al. matrices did not differentiate between commercial, recreational, and other mixing. To mitigate these concerns we averaged the mixing matrices from both sources.

States typically apply NPI and vaccination policies to specific groups. To ensure that we could represent state policies, we transformed these averaged matrices to represent the nine non-aggregated strata shown in table 3. Creating different matrices for each state would be time-consuming and unnecessary for the analyses we aim to perform with the model. Instead, we created a single set of matrices using the US populations, though the size of each stratum is allowed to vary by state. The averaged matrices contain strata by age group and chronic condition. We aggregated these strata into three age groups (age ≤ 17, 18 ≤ age ≤ 64, age ≥ 65). To create ‘employed’ and ‘not employed’ strata we split mixing in the ‘working age 18 ≤ age ≤ 64 strata. In every mixing mode except work, we split working-age mixing in proportion to the population who were employed or not employed. This is equivalent to assuming that average levels of mixing in the household, school, commercial, recreational, and other settings are the same for employed and unemployed individuals. Work mode mixing was assigned entirely to the employed strata. Based on BLS data [24] approximately 10% of workers are younger than 20 or older than 65, these workers and their work mixing was aggregated into their respective age groups.

**Table 3:** Model strata and corresponding populations

| Non-aggregated Strata | US Unique Population (millions) | Aggregated Strata |
| --- | --- | --- |
| Young | 73.4 | Young |
| FLEW without high-risk conditions | 36.0 | FLEW |
| Employed, but not FLEW, without high-risk conditions | 60.0 | Working age |
| Not employed without high-risk conditions | 33.2 | Working age |
| Old without high-risk conditions | 11.8 | Old |
| FLEW with high-risk conditions | 14.1 | FLEW |
| Employed, but not FLEW, with high-risk conditions | 37.6 | Working age with high-risk conditions |
| Not employed with high-risk conditions | 20.8 | Working age with high-risk conditions |
| Old with high-risk conditions | 37.6 | Old |

We further split the employed strata into Front-line essential workers (FLEW) and other workers. All mixing was split in proportion to population, equivalent to assuming that FLEW and other workers have similar mixing patterns. We assumed that FLEW would only mix with other FLEW at work and, similarly, that non-FLEW workers would only mix with other non-FLEW workers. This was based on the Cybersecurity & Infrastructure Security Agency (CISA) guidance which indicates that most essential industries are composed entirely of essential workers [25]. We assumed that those workers with chronic high-risk conditions have mixing behaviors identical to their colleagues without high-risk conditions.

Some of these assumptions may not be robust. For instance, essential workers may have different distributions of mixing compared to other workers in settings outside work. They may mix with other essential workers more in social settings because personal relationships are linked to work relationships and many social networks exhibit homophily [26]. Frontline essential workers may also have different levels of mixing at work. Many essential jobs, such as grocery store workers or first-responders involve high-levels of contact. However white-collar jobs are not defined by low mixing, offices and group meetings are high-contact environments, but by the ease at which mixing can be reduced during a pandemic. In an analysis of O*NET data [27], RAND colleagues found that white-collar jobs tended to have higher scores on questions that indicated higher work mixing than blue-collar or service jobs. These mixing matrices represent baseline mixing, differences in adaptability are modeled through strata-specific effects of NPIs.

Detailed strata enable us to quickly adapt if states change their vaccination policies. However, model calibration and run-times are significantly longer with more strata, especially if the strata are small (due to absolute solver tolerances). To ensure that no stratum was less than 5% of the population, we aggregated the nine strata into five, as shown in Table 3. We aggregated strata that had the same vaccination priority, for instance, FLEW are always vaccinated early irrespective of whether they have a chronic condition. Where strata were aggregated, their mixing and disease severity are the population-weighted average.

### B.2 Model parameters

Parameters indicate how people move between states in the model. These include disease progression rates, the proportion of individuals who enter more severe disease states, and the relative infectivity of each stage.

Parameter estimates were selected from a review of the literature and with the input of RAND experts. To carry out sensitivity analyses of the parameters and to calibrate the model, we constructed a large set of independent case runs, each with a different and unique combination of model parameter values. Parameter values for the case runs are sampled using a Latin-Hypercube approach [28, 29]. We use either a uniform or a beta-PERT (Program Evaluation and Review Technique) distribution to sample the model parameter value, as specified within a sensitivity analysis range [30]. In the latter case, the reference value is used to specify the mode of the beta distribution used for our parameter value sampling. The model formulas shown are based on the parameters in Figures 4, 5, and 6. Summaries of parameter estimates, sources, and sensitivity are shown in Tables 4, 5, 6 and 7

**Table 4:**
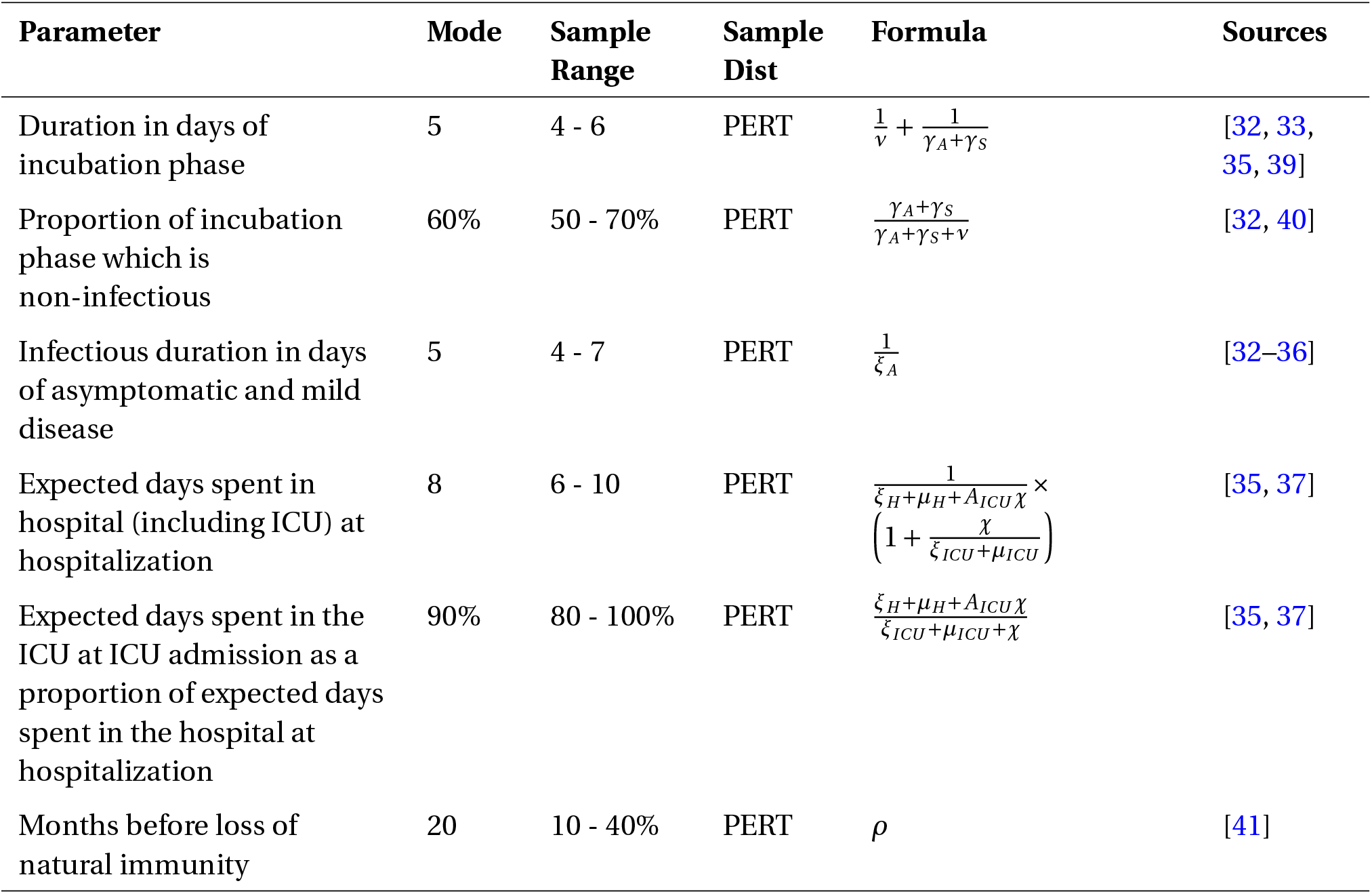
Disease duration parameter estimates

| Parameter | Mode | Sample Range | Sample Dist | Formula | Sources |
| --- | --- | --- | --- | --- | --- |
| Duration in days of incubation phase | 5 | 4 - 6 | PERT | $\frac{1}{v} + \frac{1}{\gamma_A + \gamma_S}$ | [32, 33, 35, 39] |
| Proportion of incubation phase which is non-infectious | 60% | 50 - 70% | PERT | $\frac{\gamma_A + \gamma_S}{\gamma_A + \gamma_S + v}$ | [32, 40] |
| Infectious duration in days of asymptomatic and mild disease | 5 | 4 - 7 | PERT | $\frac{1}{\xi_A}$ | [32–36] |
| Expected days spent in hospital (including ICU) at hospitalization | 8 | 6 - 10 | PERT | $\frac{1}{\xi_H + \mu_H + A_{ICU}\chi} \times \left(1 + \frac{\chi}{\xi_{ICU} + \mu_{ICU}}\right)$ | [35, 37] |
| Expected days spent in the ICU at ICU admission as a proportion of expected days spent in the hospital at hospitalization | 90% | 80 - 100% | PERT | $\frac{\xi_H + \mu_H + A_{ICU}\chi}{\xi_{ICU} + \mu_{ICU} + \chi}$ | [35, 37] |
| Months before loss of natural immunity | 20 | 10 - 40% | PERT | $\rho$ | [41] |

**Table 5:**
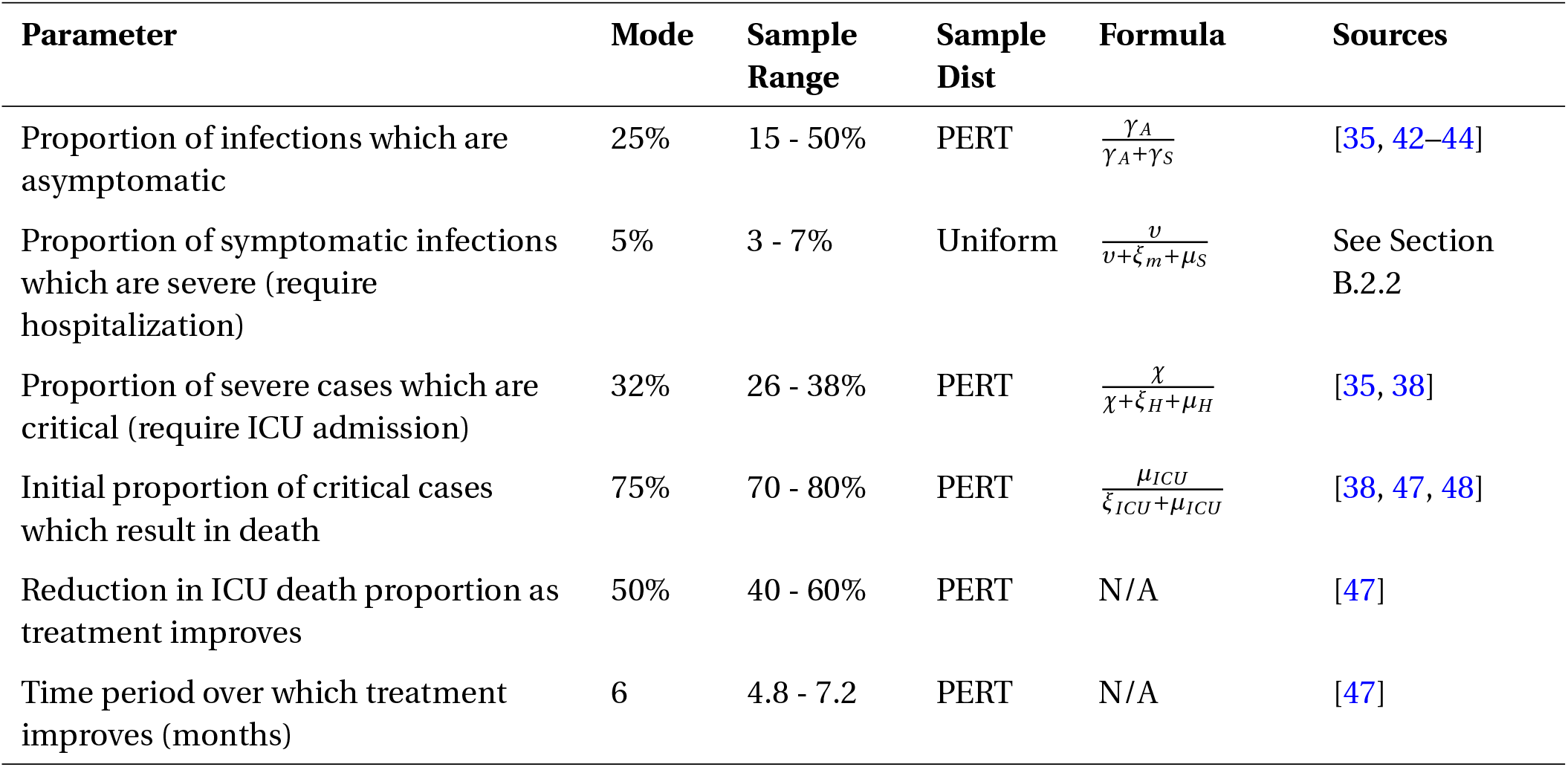
Disease prognosis parameter estimates

**Table 6:** Relative infectivity parameter estimates

| Infectivity of phase relative to asymptomatic phase | Mode | Sample Range | Sample Dist | Sources |
| --- | --- | --- | --- | --- |
| Pre-symptomatic infectious | 2.07 | 1.65 - 2.48 | PERT | [10, 31, 35, 50] |
| Mild symptomatic | 0.83 | 0.66 - 1.00 | PERT | [10, 31, 35, 50] |
| Severe symptomatic | 0.14 | 0.11 - 0.17 | PERT | [10, 31, 35, 50] |
| Hospitalized | 0.07 | 0.06 - 0.08 | PERT | [10, 31, 35, 50] |
| Tested mild symptomatic | 0.28 | 0.22 - 0.33 | PERT | [10, 31, 35, 50] |
| Tested severe symptomatic | 0.14 | 0.11 - 0.17 | PERT | [10, 31, 35, 50] |
| Tested asymptomatic | 0.20 | 0.16 - 0.24 | PERT | [10, 31, 35, 50] |

**Table 7:**
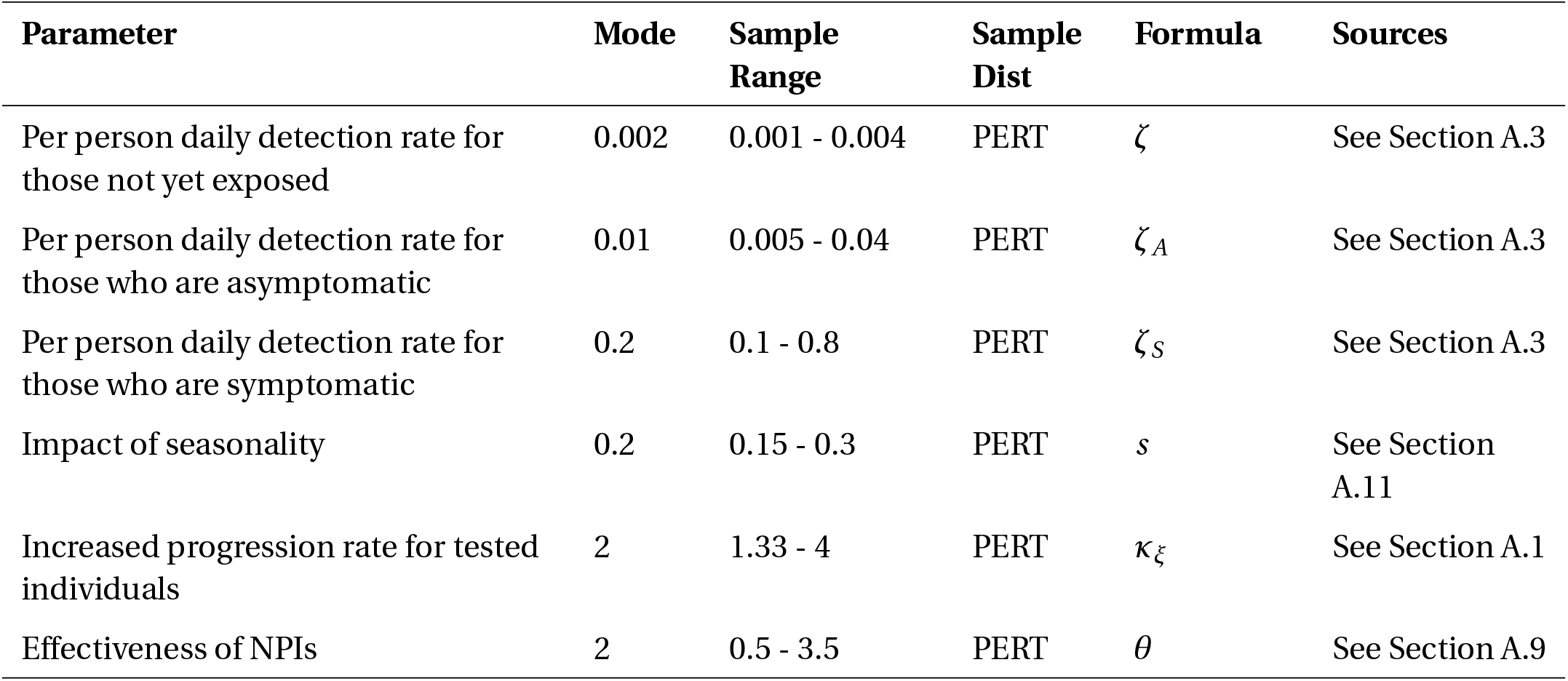
Other parameter estimates

| Parameter | Mode | Sample Range | Sample Dist | Formula | Sources |
| --- | --- | --- | --- | --- | --- |
| Per person daily detection rate for those not yet exposed | 0.002 | 0.001 - 0.004 | PERT | $\zeta$ | See Section A.3 |
| Per person daily detection rate for those who are asymptomatic | 0.01 | 0.005 - 0.04 | PERT | $\zeta_A$ | See Section A.3 |
| Per person daily detection rate for those who are symptomatic | 0.2 | 0.1 - 0.8 | PERT | $\zeta_S$ | See Section A.3 |
| Impact of seasonality | 0.2 | 0.15 - 0.3 | PERT | $s$ | See Section A.11 |
| Increased progression rate for tested individuals | 2 | 1.33 - 4 | PERT | $\kappa_\xi$ | See Section A.1 |
| Effectiveness of NPIs | 2 | 0.5 - 3.5 | PERT | $\theta$ | See Section A.9 |

**Table 8:** Shortened parameter labels of the most significant parameters found by the sensitivity analyses.

| **Parameter** | **Denoted** |
| --- | --- |
| Proportion of incubation phase which is non-infectious | Proportion Non- infectious Incubation |
| Proportion of symptomatic infections which are severe (require hospitalization) | Proportion Severe |
| Proportion of severe cases which are critical (require ICU admission) | Proportion Critical |
| Proportion of infections which are asymptomatic | Proportion Asymptomatic |
| Initial proportion of critical cases which result in death | Proportion die in the ICU |
| Duration of incubation phase | Incubation Days |
| Expected time spent in hospital (including ICU) at hospitalization | Hospitalized Days |
| Infectious duration of asymptomatic and mild disease | Symptomatic Mild Days |
| Per person daily detection rate for those who are asymptomatic | $\zeta_A$ |
| Per person daily detection rate for those who are symptomatic | $\zeta_S$ |
| Infectivity of phase of the mild symptomatic relative to asymptomatic phase | $m_{Sm}$ |
| Infectivity of phase of the severe symptomatic relative to asymptomatic phase | $m_{Ss}$ |

#### B.2.1 Duration parameters

Duration parameter estimates are shown in Table 4 and specify how fast individuals advance through the disease phases. Estimates are for the mean duration of the phase length, rather than for any individual’s phase lengths. The incubation phase, or pre-symptomatic phase, is the time from exposure to the virus to the appearance of the first symptoms. We assume that immediately after exposure individuals are not infectious, but that they become infectious before exhibiting symptoms. Some research suggests that the pre-symptomatic phase is the most infectious period [31], we explore this more in Section B.2.3. We assume that the infectious periods are the same length for those with mild disease or who are asymptomatic [31]. As shown in Figure 6 all individuals who develop severe disease must first pass through the mild disease phase. The mean duration of mild disease is similar to the mean delay between symptom onset and hospitalization [32–37], so we assume that individuals are admitted almost immediately to the hospital after entering the severe compartment of the model.

We describe hospital and ICU stays through two parameters. The first includes the expected time spent in the hospital at hospitalization, which includes some expectation of ICU admission. The second parameter is the expected time spent in the ICU at ICU admission as a fraction of the first parameter. For COVID-19 a significant proportion do not enter the ICU and ICU stays are long [37, 38], so the second parameter is a large fraction of the first. The parameters are constructed in this manner so that the length of ICU and hospital stays are not independently sampled.

#### B.2.2 Prognosis parameters

Prognosis parameter estimates are shown in Table 5 and specify what fraction of individuals enter different phases, such as whether individuals with mild disease recover or develop severe disease. We define severe disease as requiring hospitalization and critical disease as requiring ICU admission. The proportion of asymptomatic individuals used in this model is substantially lower than early estimates. Recent systematic reviews have found that many early studies overestimated the fraction of symptomatic individuals because they applied restrictive criteria for symptoms or did not observe participants for long enough to determine if they were asymptomatic or presymptomatic [42–44].

The proportion of symptomatic infections which result in hospitalization has not been reliably estimated in the literature. Available estimates find high rates (19%) but are likely biased due to selection effects which undersample mild cases [45]. Merely looking at the ratio of cumulative hospitalizations to cumulative positive tests in US (among states who report both metrics) as of February 2020 [46] shows a symptomatic hospitalization rate of around 6% (accounting for the proportion symptomatic shown in Table 4). The real figure is likely significantly lower than this number because we observe a higher fraction of hospitalizations due to COVID-19 than we do COVID-19 infections.

There is evidence that the fatality rate of those hospitalized has radically decreased since the beginning of the pandemic due to a combination of improved protocols and new treatment regimens. Part of this effect is explained by a change in the age composition of the hospitalized, but accounting for this some researchers estimate a 2 to 3-fold decrease in fatality rate among those admitted to the hospital [47]. This finding suggests that treatment efficacy is an important time-varying confounder in our model. Therefore, we allow a treatment efficacy stock variable *T* that is initiated at zero is modeled as *d T* /*dt* = (*T* ^*^−*T*)/*t*_*p*_ where *T* is the treatment efficacy, *T* ^*^ is the final treatment efficacy, and *t*_*p*_ controls how fast treatment improves. To model this, we assume that a high proportion of those admitted to the ICU die. This formulation allows treatment efficacy to become a time-varying variable, as it has been observed empirically [47].

#### B.2.3 Relative Infectivity parameters

The relative infectivity, the daily expected infections of susceptible, varies by disease state. We conceptualize infectivity as having two components: contact mixing, how likely people are to mix with others, and biological transmissibility, the probability of transmission between an infectious and a susceptible person given a contact. The infectivity is the product of these components. We define infectivities relative to the asymptomatic state.

To calculate biological transmissibility we rely on viral load data and estimates of the number of infections in the presymptomatic period. We use viral load data from individuals who tested positive [31], assuming that the average test occurs 3 days after initial exposure [35]. Based on relationships between biological transmissibility observed in other infectious diseases, we assume that the functional form for the relative biological transmissibility, *β*_1,2_ between the viral load in two states, *V L*_1_ and *V L*_2_ is:

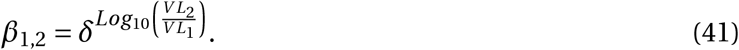

Where *δ* is a calibration parameter. Several studies have estimated that the fraction of infections caused during the presymptomatic period is approximately 45% [10, 49]. We choose *δ* = 1.92 such that 45% of infections are caused in the presymptomatic phase once contact mixing is accounted for. We also assume that those who are asymptomatic have biological transmissibility only 75% of those who have mild symptoms based on estimates by the CDC [36]

Contact mixing changes by disease phase because individuals may be incapacitated COVID-19 or may voluntarily reduce their mixing to avoid infecting others. We assume that those who are presymptomatic or asymptomatic do not modify their mixing relative to those who have not yet been exposed to the virus. Those with mild symptoms reduce mixing by 40%, those who are tested positive by 80%, those who are severe (but not yet hospitalized) by 90%, and those who are hospitalized by 95%. The reduction for those who are hospitalized is larger than for those who are waiting for hospitalization because hospital staff is expected to have better protective equipment than home carers. The infectivities relative to the asymptomatic phase are shown in Table 6.

#### B.2.4 Other parameters

We randomly sample several other parameters from distributions. These are shown in Table 7 and pertain to testing rates, seasonality, and the effectiveness of NPIs within that state. See Sections A.3, A.9, and A.11 for more details.

##### Testing and Detection Rates

In our most recent model runs, we do not assume constraints on the availability tests. Hence we assume constant rates for *ζ, ζ*_*S*_, and *ζ*_*A*_. During December 2020 and January 2021, the total daily rate of tests in the US ranged between 1.5 and 2 million tests a day, and the positive rate was 10%. We assume the infectious duration of the mild disease is between 4 and 7 days, and between 33% and 66% of the mildly symptomatic infected individuals seek to get tested during this time. Using these numbers, we estimate that the per-person detection rate of the mildly symptomatic *ζ*_*S*_ ranges between 0.06 to 0.13 per day. Using the 10% positive rate and the infectious duration of asymptomatic disease ranging between 4 and 7 days, we estimate that the per-person detection rate of the asymptomatic *ζ*_*A*_ ranges between 0.01 and 0.012 per day.

##### Seasonality

The range of values used for our seasonality parameter *s* was based on estimates used for influenza [51]. However, depending on the climate zone, the range of *s* varied from 0.3 to 10. A more recent study that looked at seasonality patterns for two human coronaviruses (CoVHKU1 and CoVOC43) found a much lower and tighter range [52]. Based on this study, we assume that *s* varies between 0.15 and 0.3.

##### Increased progression rate for tested individuals

We estimate that, on average, those who tested with mild symptoms will have experienced symptoms for a few days before being diagnosed. Hence they are more likely to progress to the next disease state than those that do not seek a test. We estimate that the time they have already spent with mild-symptoms is between 25% and 75% of the expected duration of experiencing mild symptoms. Inverting these proportions, we estimate that the range for *κ*_*ξ*_ is between 1.33 and 4 with a most likely value of 2. Similar reasoning and estimation apply to the asymptomatic and how long they are infectious before seeking to be tested if they suspect to have had contact with a confirmed case.

## C Sensitivity Analysis

This section presents a sensitivity analysis of the model runs generated by an experimental design that considered 2,000 independent cases, each with a unique combination of input parameter values generated by a Latin Hypercube Sampling (LHS) approach. Each case run considers the COVID-19 dynamics for California from March 1st, 2020, to December 31st, 2020. This is the period of interest before the distribution of the vaccines. Because our period of interest has a duration of less than a year, we fix some model parameter values to their mode value. We do not consider them in the sensitivity analysis. Most notably, we fix the loss of immunity rate *ρ*.

We focus on the three model outputs produced. These are the cumulative deaths and the cumulative cases diagnosed by the end of 2020, and the hospitalized cases on that date. We use two approaches. The first is to compute the partial rank correlation coefficient (PRCC) [29] and the second is based on Classification And Regression Tree (CART) analysis.

The partial correlation between model output and a model input measures the degree of association between the two while controlling for the variability and the effects of the other model inputs. The partial correlation coefficient removes issues between confounding inputs. Therefore, it is generally a better measure than simple correlation and can reveal an association between an input and an output that is the inverse of that suggested by a simple correlation.

Figure 7 shows the PRCC values associated with the input parameters. Parameters that have been omitted from the plot have negligible values.

**Figure 7:**
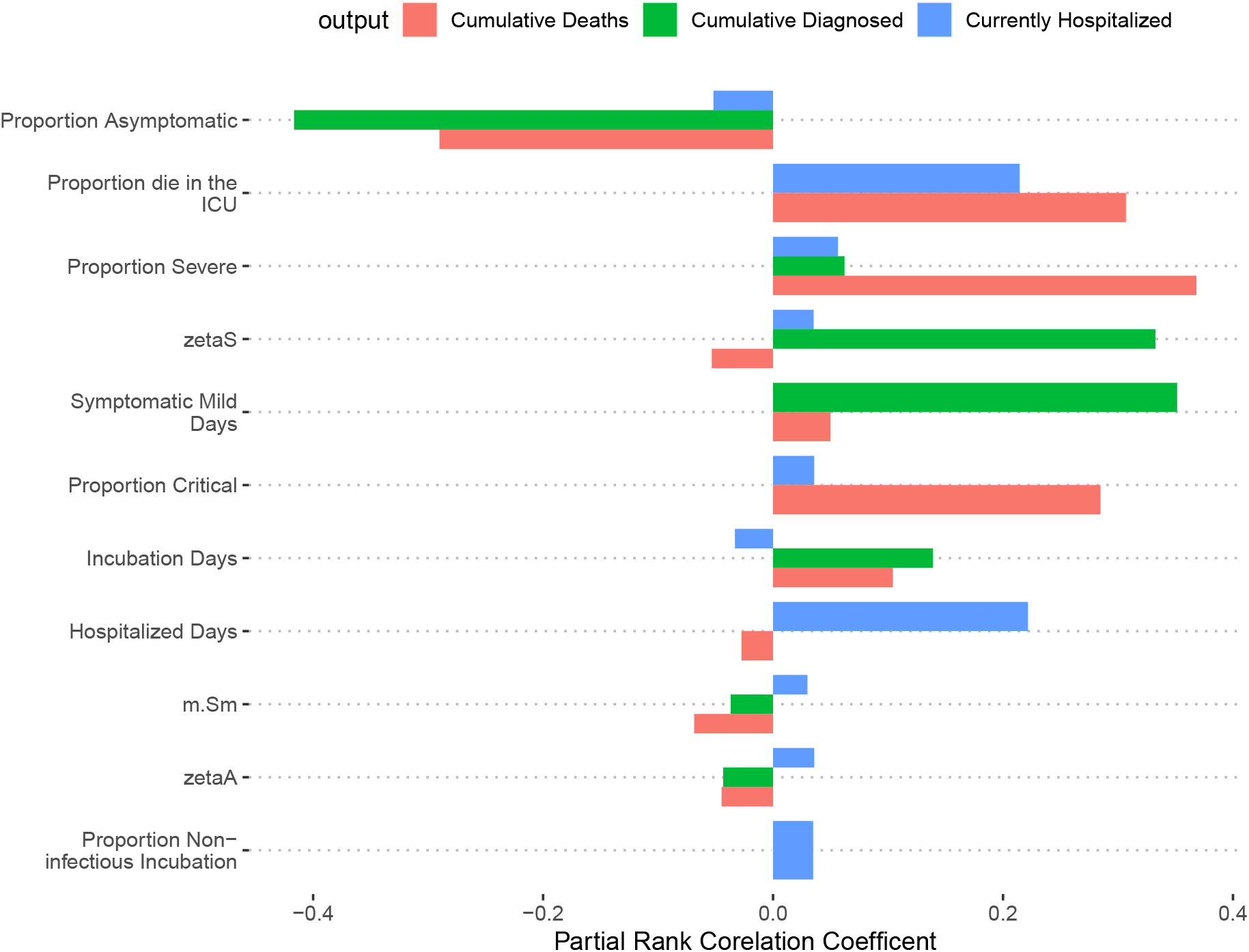
Partial rank correlation coefficient. values associate with each input parameter for our three model outputs,

Our second method uses the importance scores of a CART algorithm. CART is a supervised machine learning algorithm that is commonly applied to analyze large data sets [53, 54]. For each model output of interest, CART creates a binary decision tree. This process recursively splits the space of input parameter values into regions that produce comparable output. As a first step, the algorithm uses regressions to find which input parameter and its range in values best explain the model output variance across all the model case runs. Using a cost function, it finds the input parameter’s threshold value that best splits the output data into two distinct groups. Using the CART terminology, this creates the root-node and two first-generation leaf-nodes. If the input parameter for splitting is numerical and continuous, then the algorithm uses a regression-based method. If the input parameter is categorical, then a different approach is used to use a Gini index function instead of a cost function. This index indicates how “pure” a leaf-node is in terms of what different categorical values of the input it includes. During this process, all input parameters and all possible split point values are evaluated. For a given input parameter and split point, the algorithm first splits the data into two separate training samples and runs a regression model for each sub-sample. For each of the two sub-samples, it then uses the sum of the squared difference between the training sub-sample data and the regression’s prediction to generate the cost function. The algorithm then chooses the input parameter and the split point that minimizes the cost function. Thus, the cost function is minimized when the input space is split into two regions that produce distinct outputs.

The algorithm repeats this process for each of the two first-generation groups or leaf-nodes. Thus, using the subset of model case runs belonging to each group, we repeat the process and find which input parameters can best split the output data into additional second-generation leaf-nodes. This process continues until a stopping criterion is satisfied. The most common stopping procedure is to specify a minimum number of model case runs that need to be assigned to each leaf node. When the method splits the model cases runs belonging to a given node into two leaf-nodes, and either leaf-node has fewer cases than some minimum threshold number, then the split is not accepted, and the node is taken as a final leaf-node. Alternatively, stopping criteria can be specified based on when the variance of the output across all model cases contained a given leaf-node goes below a specific threshold value.

The CART plots shown in figures 8 - 10 show the decision tree for each of the three model outputs and help provide a visual intuition of the importance of each input parameter in splitting the data.

**Figure 8:**
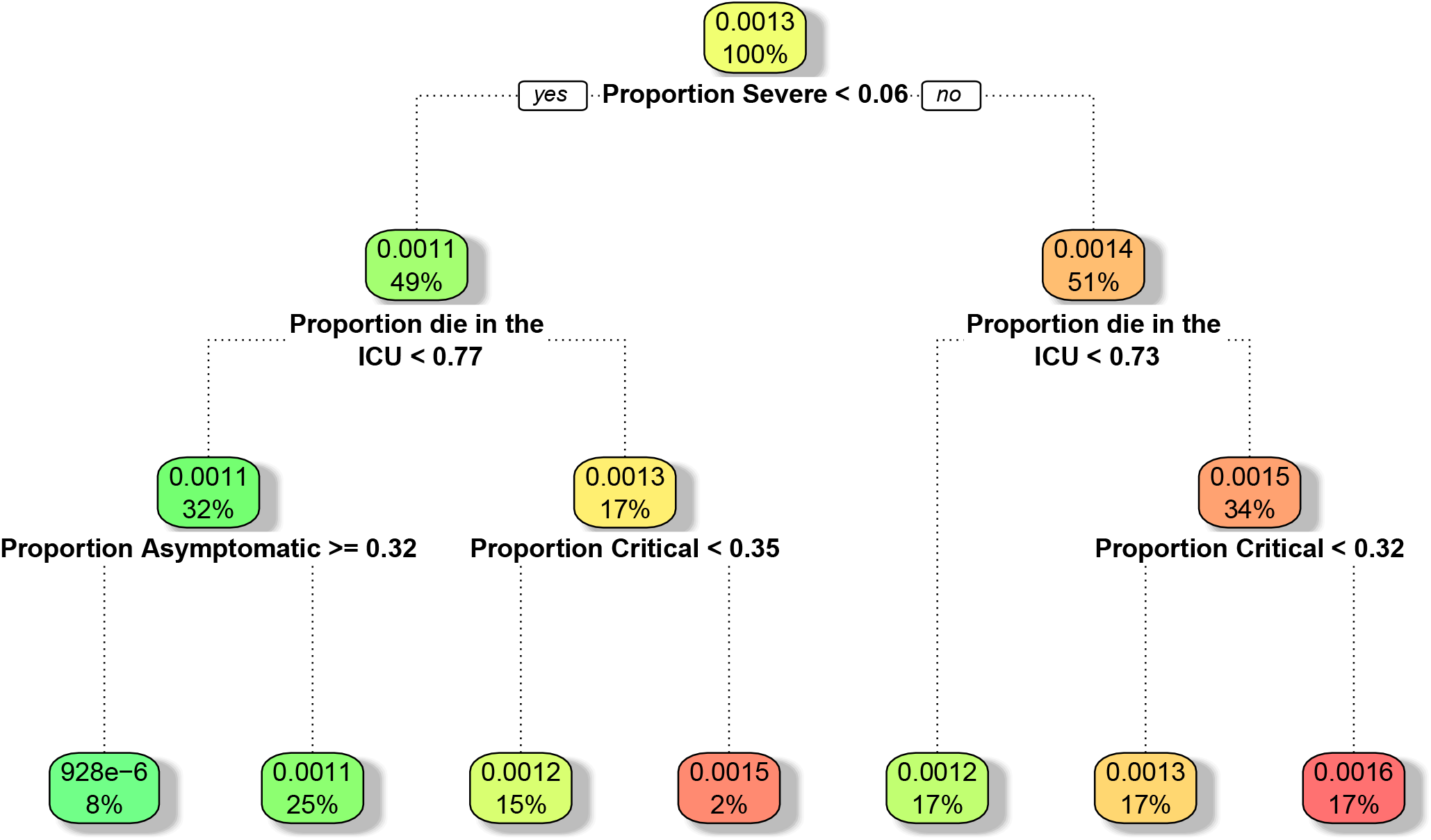
Cumulative Deaths CART decision tree.

**Figure 9:**
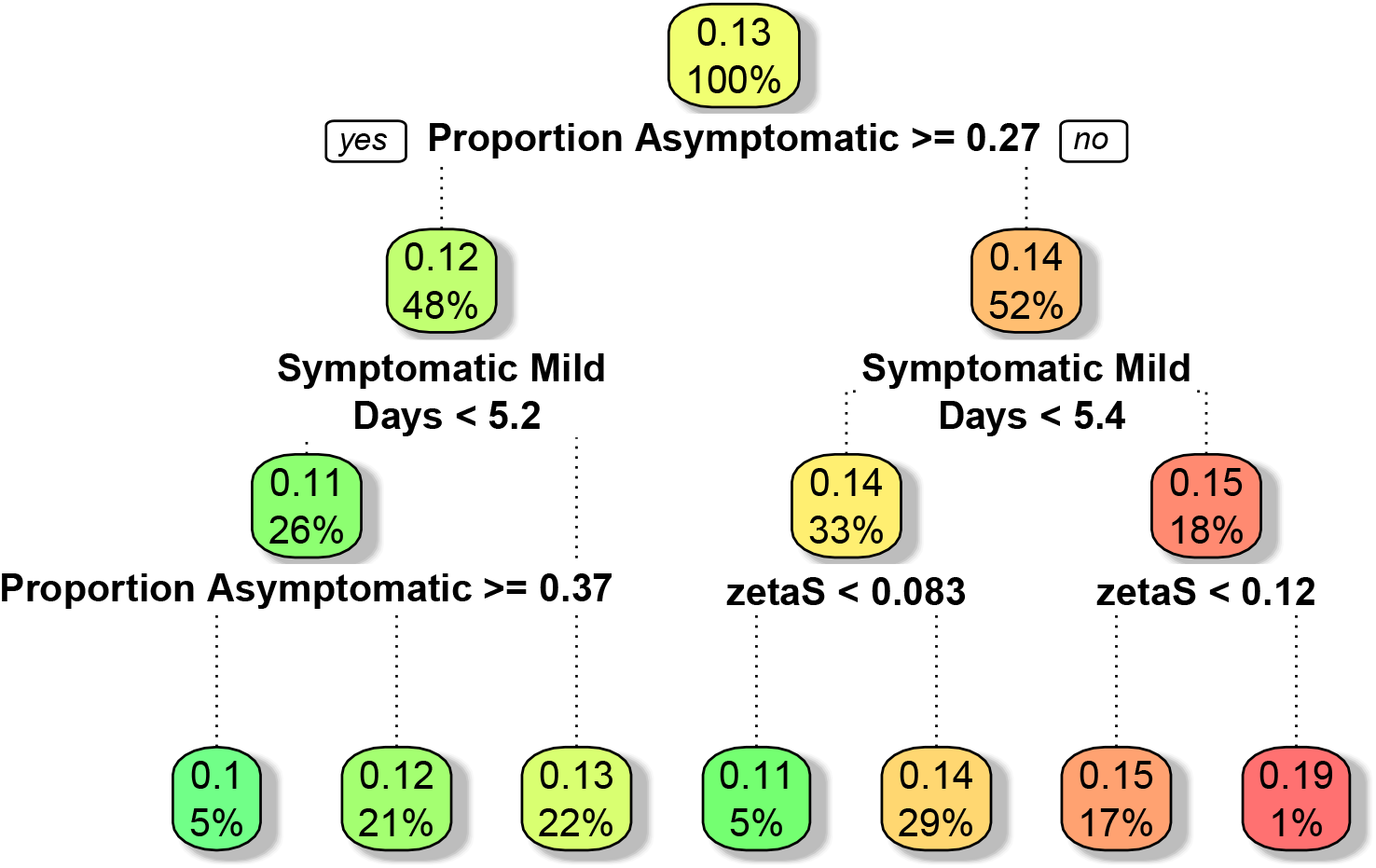
Cumulative Diagnosed CART decision tree.

**Figure 10:**
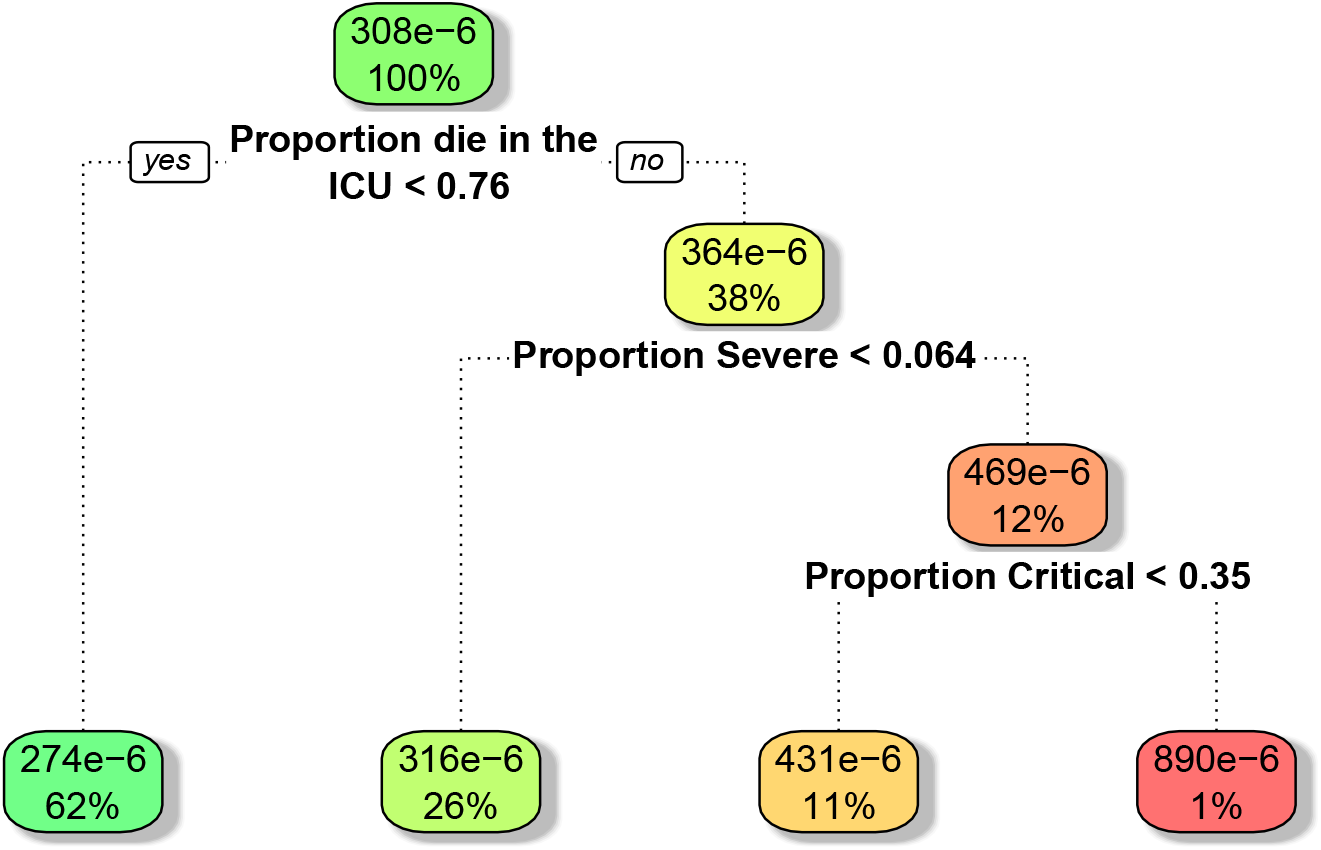
Hospitalized CART decision tree.

An overall importance score can be associated with each node calculated over the CART procedures. The importance score is related to how often each input parameter is used to split the output data most optimally over the decision tree’s nodes. It also considers how well the less optimal parameters split the data at each node without the more optimal parameters. Optimally at each node is found by finding the improvement scores associated with each node and measuring the increase in the split’s quality in the data. The parameter with the highest quality best breaks the data into two distinct output sets. The importance score takes into account which generation the node belongs to for the split. Parameters that split the data at the root node or the first-generation node have more weight. Parameters used in later generation data splits carry less weight towards calculating their importance score even if they produce high-quality data splits.

Figure 11 shows a plot of the CART importance scores associated with each input parameter.

**Figure 11:**
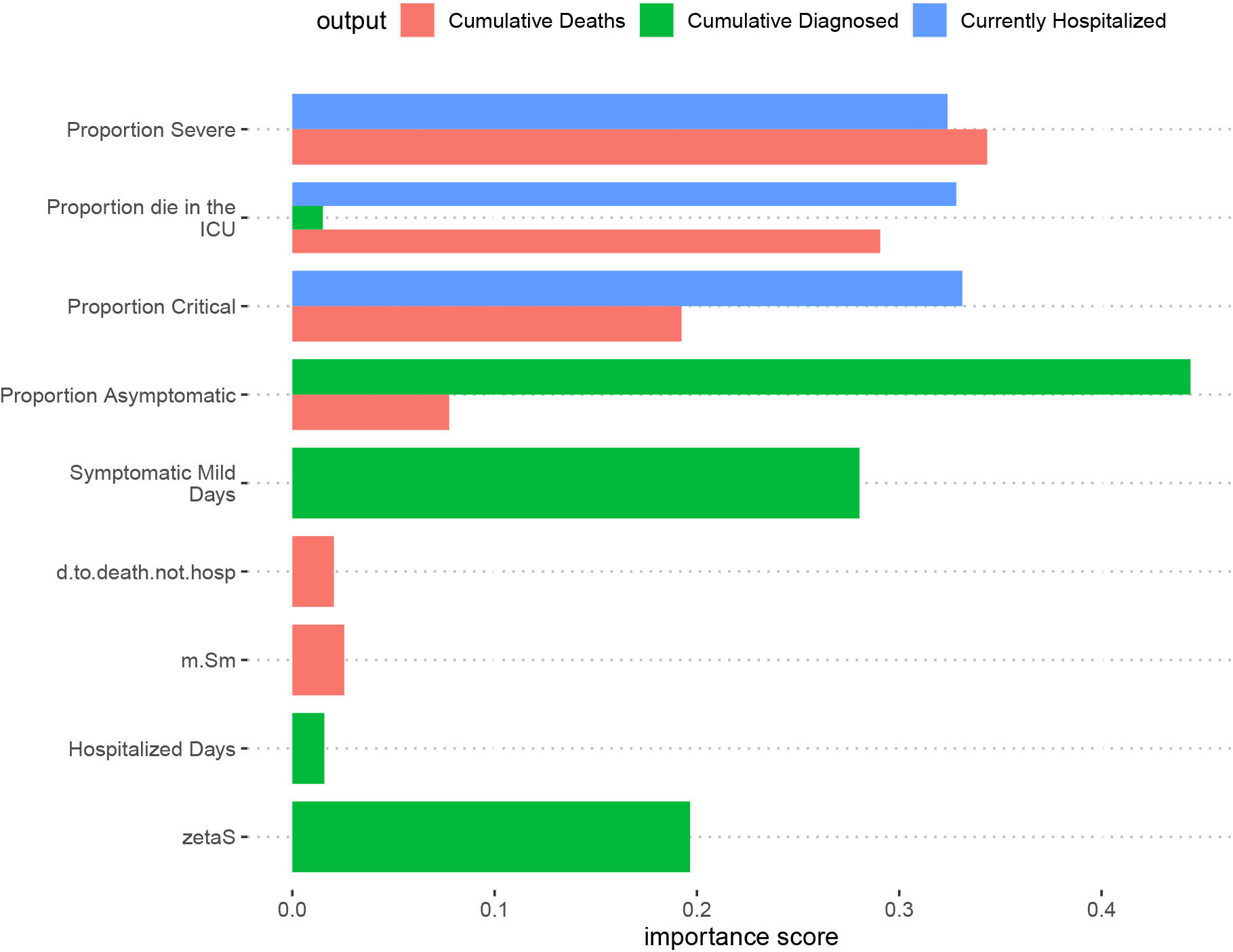
CART Importance scores for each model outputs and associated with each input parameter.

## D Limitations

Population-level models are best suited to comparing potential interventions, rather than to forecast outcomes. They make strong assumptions about functional forms, individual behavior, and how parameters (such as treatment efficacy) change over time. These assumptions allow them to be informed with relatively little data but limit the extent to which they can reliably reproduce or predict the outbreak [55]. In this section, we detail several other model limitations.

### Unobservable True Cases

Ideally, we would observe the true case count. However because cases can only be verified through testing, the true case count is not observed, instead, we use proxies: confirmed cases and deaths. Both these proxies have flaws. Observed cases are confounded by testing rates, increases in test availability have caused surges in confirmed cases leading to the overestimation of *R*_0_. Deaths are also an imperfect proxy due to the delay between infection and death. Deaths may also be confounded by testing in states where regulations require a positive test to report deaths.

### State Mixing

The model assumes that no mixing occurs across state lines. This assumption is of little importance while the virus remains widespread in almost every county in the United States. However, as the virus becomes more controlled in some areas of the country, travel bans may become an important tool to stop reseeding events.

### Behavioral Feedback

To make accurate predictions about the future or counterfactuals about the past, one needs to understand how populations will react to different circumstances. Compliance is crucial to the effectiveness of NPIs but is likely a function of perceived risk. As cases decrease, even if mandated NPIs remain stringent, compliance may decrease to the point where we are unable to contain the spread of the virus. Conversely, people may limit their mixing voluntarily if they perceive significant personal risks. Our PBM does not contain an endogenous model of mixing behavior and so has limited ability to model the second-order effects in counterfactual scenarios.

## Footnotes

1 One might estimate the costs of NPIs using a willingness to pay or similar approach. As long as those weights are monotonically increasing (e.g. people are not deriving utility from NPI restrictions), our substantive findings would hold. While estimating more precise welfare costs of NPIs and using those costs as weights might be valuable to compare benefits from NPIs to costs, we doubt that these weights would be stable over time. Still, as long as these weights are monotonically increasing functions of the NPI level *at any point in time* our substantive results would hold. Because these weights are highly uncertain and potentially not constant, we refrain from trying to aggregate all outcomes under a single social welfare metric in our analyses as a traditional Cost-Benefit analysis would do. Instead, we assess pareto-efficiency and seek to find strategies that dominate other strategies across a set of outcomes.

